# How have restaurant menus changed following England’s calorie labelling regulations and who is likely to benefit? A longitudinal analysis of online menu data

**DOI:** 10.64898/2026.01.27.26344846

**Authors:** A Kalbus, R Kumar, C Rinaldi, E Curtin, J King, P Reynolds, L Cornelsen, M Essman

## Abstract

**Background:** The introduction of mandatory calorie labelling among large food businesses (chains) in England in 2022 has been found to have little impact on consumer behaviour, but overall calories on restaurant menus have decreased slightly. This study examined menu changes post policy implementation, and the population groups likely to have been affected most.

**Methods:** Menu data from 169 chains in Great Britain were extracted from two online food delivery platforms in June 2022 and June 2023. We selected 10 categories (specific foods or chains) jointly with public and policy advisors. Menu changes over time were assessed with multilevel models accounting for whether an item was continuously on the menu and for the type of chain. Where changes were found, we assessed differences in purchasing frequency by consumer characteristics using 2022 OOH purchase data (Worldpanel by Numerator, GB OOH Panel).

**Results:** Changes were observed in two (out of 10) categories examined and were driven by changing items on the menu rather than reformulating continuous dishes. Chains that used a ‘healthy’ tag on the delivery website increased the share of mains under 600 kcal by 3.7 percentage points (95% CI 0.2 to 7.2), while average calories did not change (-17.6 kcal/item, 95% CI -38.7 to 3.4). Men, people aged 35–44 years and with high SES were found to purchase more frequently from these chains. Across all chains, the share of lower-calorie coffees decreased by 10 pp (95% CI -18.0 to - 0.02), with purchasing more frequent among men and increasing with age.

**Conclusions:** Although data were available for one year only post-policy implementation, menu changes among the investigated foods and chains were limited. While menu change may equitably improve population dietary health, dietary inequalities may exacerbate if only ‘healthy’ chains already offering lower-calorie food change their menus.

## Introduction

Dietary health is a major public health concern globally [1], with diet-related disease disproportionally affecting disadvantaged population groups [2]. Food prepared outside the home is of particular concern as it is both frequently consumed, with an average of 300 kcal per person per day in Great Britain [3], and of lower nutritional quality and higher calorie density compared with foods prepared at home [4,5].

As part of policy efforts targeting the out-of-home (OOH) food sector, calorie labelling regulations were implemented in England in April 2022, which require food businesses, including restaurants and takeaways, with 250 employees or more to show the calorie content of food and non-alcoholic drinks ready for immediate consumption on their menus [6]. The legislation was first announced in the Tackling Obesity Strategy [7], is similar to international policies [8], and follows on from previous voluntary strategies which achieved limited success in improving the OOH food environment [9,10].

Calorie disclosure is hypothesised to alter calorie intake via two pathways: 1) consumers may use the information to alter their food and drink choice; and 2) businesses may lower the calorie content of their menu items via reformulating existing dishes, removing high-calorie items, or introducing low-calorie items in response to the policy. Most evidence on the impact of calorie labelling focusses on the consumer-level mechanism. A recent meta-analysis, based predominantly on experimental research from the US, concluded that calorie labelling can lead to a reduction of 11 kcal per meal [11]. Real-world evaluations, on the other hand, present mixed findings [12]. In England, the recent calorie labelling policy has not been found to lead to a change calories purchased or consumed [13,14].

With respect to the second pathway, a meta-analysis of US studies found an average reduction of 15 kcal per menu item associated with calorie labelling [15]. In England, an analysis of 78 restaurant chains found a mean reduction of 9 kcal per item (95%CI -16 to -1) between September 2021 and September 2022, with some evidence of a reduction in higher-calorie items [16]. Changes in calories offered on menus vary depending on the types of food and drink offered, the menu section and restaurant type. To date, only one study investigated overall menu changes in the UK associated with calorie labelling and observed differences by chain types and broad item categories [16]. This study aims to complement the existing evidence by assessing the effectiveness and differential impacts of the calorie labelling policy in the OOH food sector in England through reformulation and menu changes. Specifically, our study (1.) estimated changes in energy content of food groups and across restaurants in selected categories; and (2.) assessed which population groups were most likely to be affected by these changes by examining purchasing by age, sex, and socio-economic status (SES). We focus on Great Britain despite the legislation pertaining to England only as many cross-border chains have implemented calorie labelling beyond England [17].

## Methods

### Design

We undertook a longitudinal analysis of menu changes in selected food groups and chains, supplemented by a cross-sectional analysis of consumer purchasing data to ascertain which population groups may be most affected by such changes. The study design is depicted in Figure 1. Supplementary Material 1 describes the public and policy involvement throughout the study.

**Figure 1.**
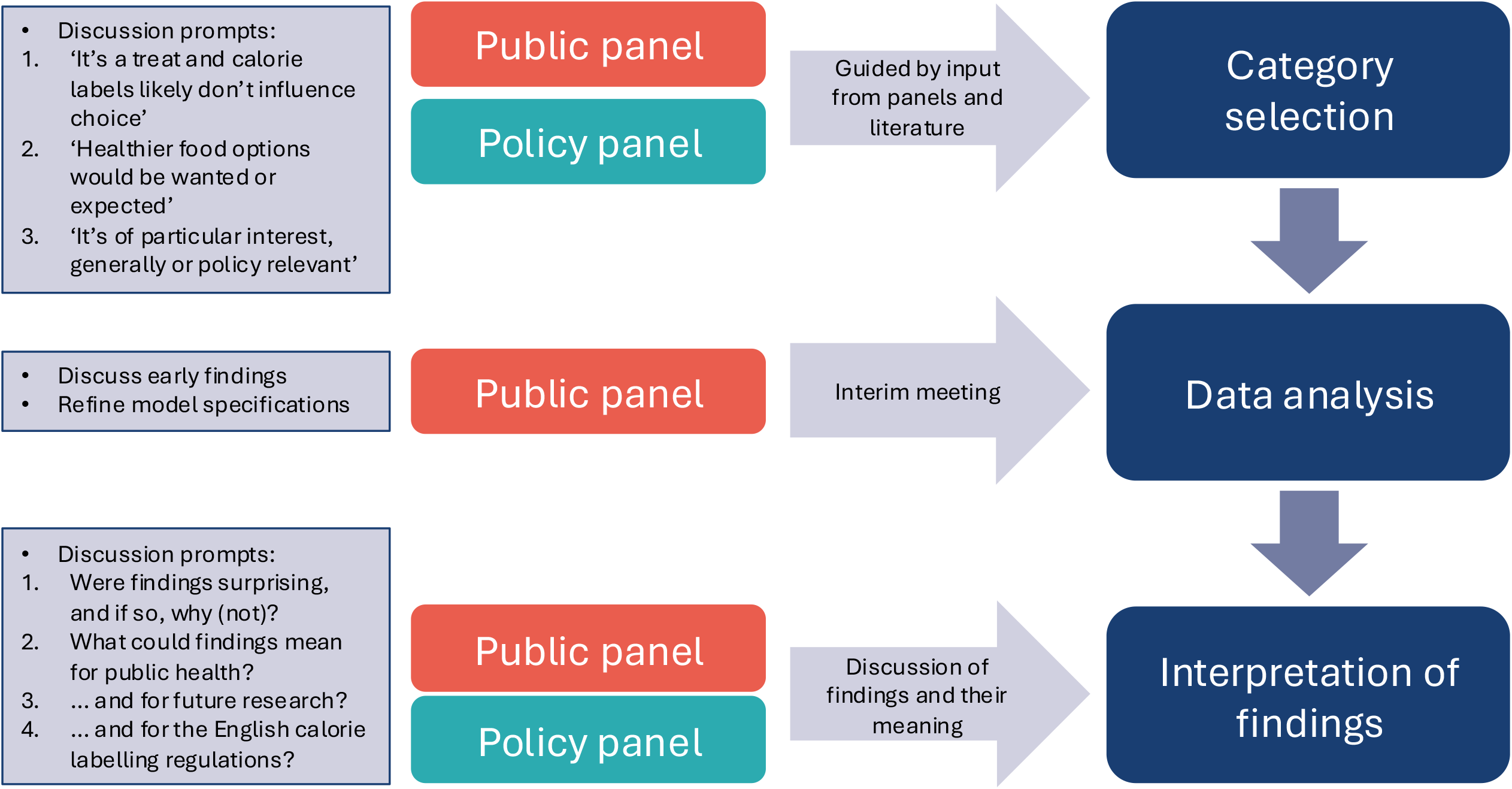
Study overview.

### Category selection

Categories to be analysed, i.e. specific foods and chains, were selected based on input from panels of public and policy advisors as well as previous literature. Sessions with the panels were structured along prompts shown in Figure 1. We included categories deemed most relevant by each panel and the literature, prioritising those raised by more than one of these three, and which could be realised using the available data (see Table 1 and Supplementary Material 1).

**TABLE 1.**
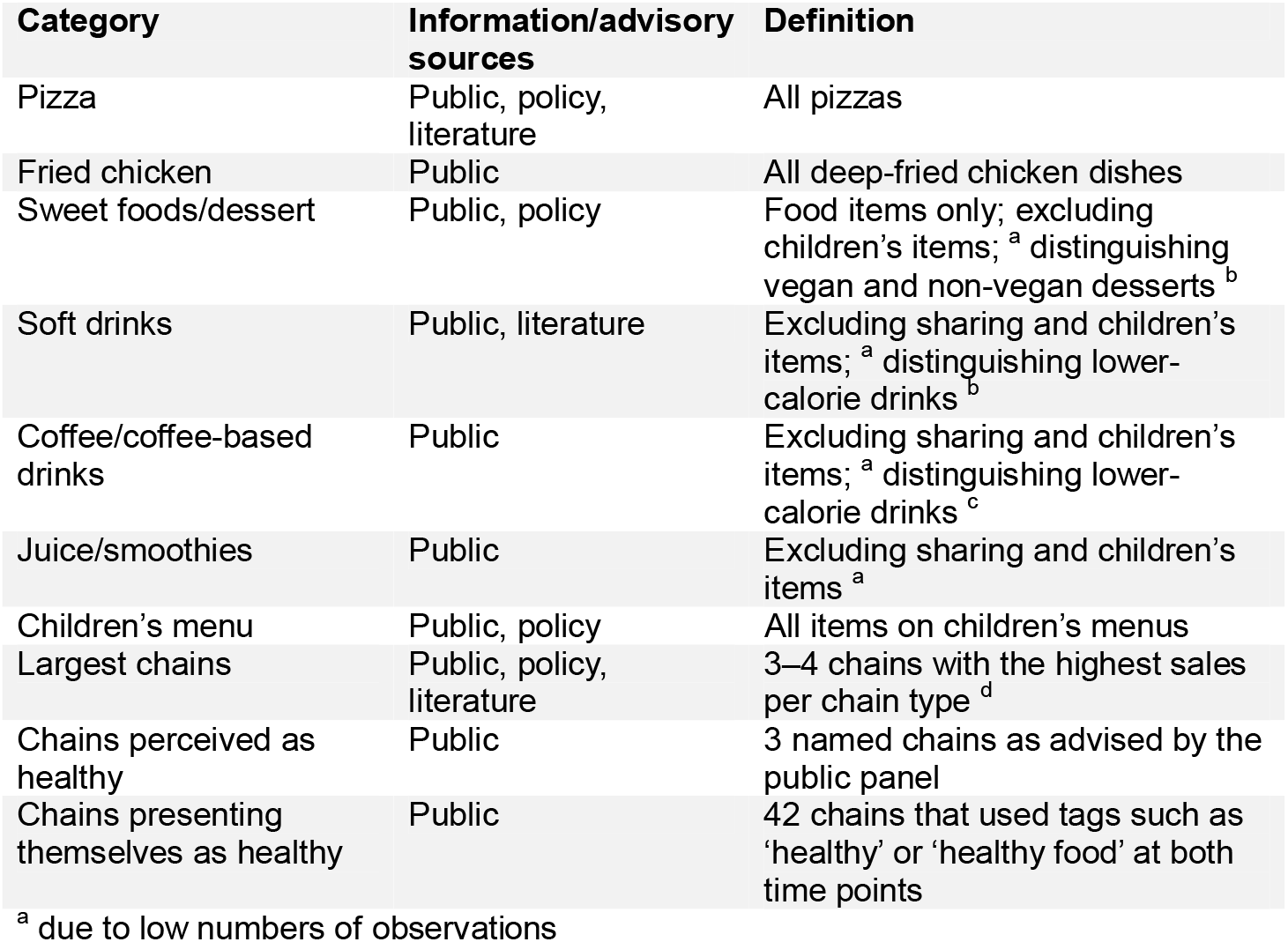
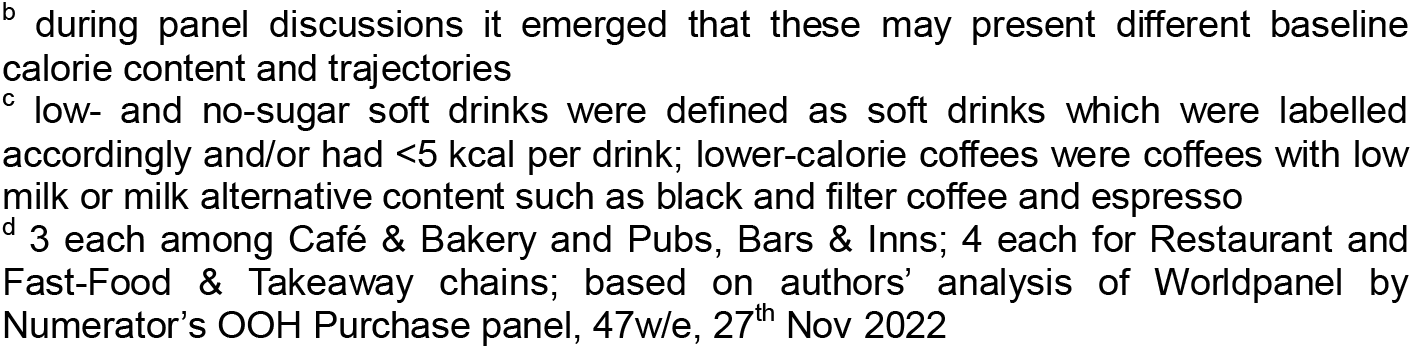
CATEGORIES.

| Category | Information/advisory sources | Definition |
| --- | --- | --- |
| Pizza | Public, policy, literature | All pizzas |
| Fried chicken | Public | All deep-fried chicken dishes |
| Sweet foods/dessert | Public, policy | Food items only; excluding children's items; <sup>a</sup> distinguishing vegan and non-vegan desserts <sup>b</sup> |
| Soft drinks | Public, literature | Excluding sharing and children's items; <sup>a</sup> distinguishing lower-calorie drinks <sup>b</sup> |
| Coffee/coffee-based drinks | Public | Excluding sharing and children's items; <sup>a</sup> distinguishing lower-calorie drinks <sup>c</sup> |
| Juice/smoothies | Public | Excluding sharing and children's items <sup>a</sup> |
| Children's menu | Public, policy | All items on children's menus |
| Largest chains | Public, policy, literature | 3–4 chains with the highest sales per chain type <sup>d</sup> |
| Chains perceived as healthy | Public | 3 named chains as advised by the public panel |
| Chains presenting themselves as healthy | Public | 42 chains that used tags such as 'healthy' or 'healthy food' at both time points |

<sup>a</sup> due to low numbers of observations
<sup>b</sup> during panel discussions it emerged that these may present different baseline calorie content and trajectories
<sup>c</sup> low- and no-sugar soft drinks were defined as soft drinks which were labelled accordingly and/or had <5 kcal per drink; lower-calorie coffees were coffees with low milk or milk alternative content such as black and filter coffee and espresso
<sup>d</sup> 3 each among Café & Bakery and Pubs, Bars & Inns; 4 each for Restaurant and Fast-Food & Takeaway chains; based on authors' analysis of Worldpanel by Numerator's OOH Purchase panel, 47w/e, 27<sup>th</sup> Nov 2022

### Data

#### Online menu data

Building upon previous work [17], custom data collection tools retrieved menu data from the websites of Deliveroo [18] and Uber Eats [19] from all food outlets delivering to each small area in Great Britain (LSOA in England and Wales and Data Zone in Scotland). We included data from June 2022, when the calorie labelling regulations had just been implemented, and one year later, in June 2023. We defined chains as businesses which were listed on MenuTracker, which contains menu data from large UK food businesses [20] and/or had ≥5 branches present in the delivery data.

We removed items without calorie information, groceries, and alcoholic drinks. To extract one representative menu per chain for each point in time, we included items that were offered by at least two branches of each chain, as determined through the item’s name. We included all items from the two chains with fewer than three branches in the data. For 8.4% of items in 2022 and 10.9% in 2023, calorie information of the same item, as determined through having the same name offered by the same chain, differed between branches. We resolved this by taking the modal calorie value for these items.

We further filtered chains included in the study as shown in Figure 2. The final dataset thus included 169 chains with 9,957 items in June 2022 and 10,158 items in June 2023.

**Figure 2.**
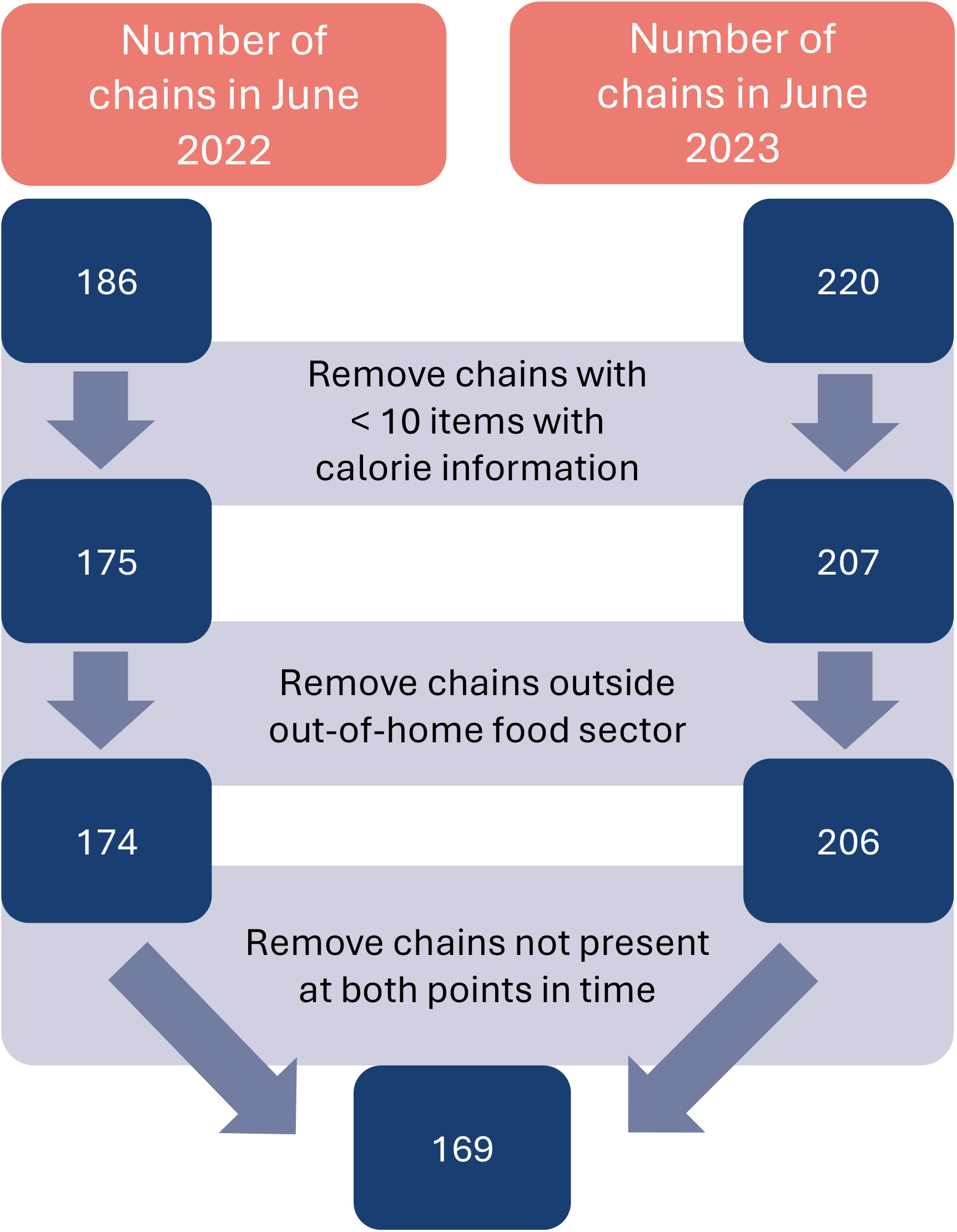
Process of filtering chains included in the study. Chains were defined as food businesses listed on the MenuTracker database [20] and/or being represented by five or more branches in the delivery data.

In line with the MenuTracker database categories [20], we classified chains as ‘Restaurants’, ‘Café & Bakery’, and ‘Pubs, Bars & Inns’, while we combined the categories ‘Western Fast Food & Takeaway’ and ‘Asian Fast Food’ to ‘Fast-Food & Takeaway’. We also determined specialty chains corresponding to the selected categories of specific foods/drinks (e.g. pizza chain, coffee shop).

We classified food and drink items according to the investigated categories (see Table 1) and determined whether an item was intended for sharing, a meal deal/bundle, or on the children’s menu through its name, description, or menu category. For chain-specific categories, we also determined whether a main food item was under 600 kcal, corresponding to government recommendations as maximum calories per lunch or dinner [21]. Based on items’ names, we distinguished those continuously on the menu (present in 2022 and 2023) from new/removed items (present at only one time-point) [16].

#### Consumer purchase data

Consumer purchase records were obtained from Worldpanel by Numerator’s Out-of-Home Purchase Panel for the period 3^rd^ January 2022 to 27^th^ November 2022. This rolling panel consists of ∼7,500 individuals representative of the population aged 13– 79 years in Great Britian with respect to age, sex, and region of residence. Panellists record all food and drink purchased for consumption away from home via a mobile phone application. Records include the item’s name and price; an identifier of the purchase occasion which denotes when several items were bought together; a store identifier, which includes names of larger retailers and generic categories for smaller businesses; and who it was bought for, e.g. for the individual themselves or for others. Information on the reporters included their age, sex, and SES. The occupation-based SES classification follows the National Readership Survey [22], and for this research was aggregated to high (AB), middle (C1C2) and low (DE).

We removed purchase records from retail and other businesses outside the OOH food sector, and considered purchases made for the individuals only, leaving 221,525 purchase occasions. We considered purchases from large chains only, which were identified by name in the data and subject to the calorie labelling regulations (n=92). Finally, we determined the frequency of purchase occasions in total and of those including each investigated category per individual.

OOH purchase recorders were predominantly female (62.3%), had middle SES (61.1%), and an average age of 48.5 years (standard deviation 15 years). Individuals reported a median of 14 OOH purchase occasions during the study period, of which just under half (46%) occurred in large chains.

### Statistical analysis

#### Research Question 1: Changes in calorie content over time

We first obtained naïve change estimates via unpaired t-tests and z tests of proportion. In an adjusted analysis, we used random intercept models to account for the hierarchical structure of items nested in chains, which themselves are nested in the type of chain. Negative binomial regression was used to model items’ calorie content and logistic regression for the prevalence of lower-calorie soft drinks and coffees as well as prevalence of main food items under 600 kcal from the largest chains, chains perceived as healthy, and chains presenting themselves as healthy over time.

Models included interaction terms between time (dummy variables for June 2022 and June 2023) and the type of chain, and between time and whether an item was continuously on the menu. Depending on the category and data availability, additional terms were included: models for pizza and fried chicken further adjusted for whether the item was on the children’s menu, a meal deal/bundle, and intended for sharing. The analysis of sweet food/desserts was also adjusted for whether the item was intended for sharing and included an interaction term between time and whether the item was vegan. Models of the calorie content of soft drinks and coffees also included a dummy variable indicating whether the item was a lower-calorie drink. The model of children’s items adjusted for whether the item was a drink or a meal deal/bundle. Finally, chain-specific categories included an interaction between time and menu section.

To assess the robustness of findings, we repeated the analysis excluding items for which multiple calorie values were recorded in the raw data.

#### Research Question 2: Differences in purchasing by sociodemographic characteristics

We examined differences in purchasing by age (in 10-year age bands), sex, and SES. We assessed absolute frequency of purchase occasions including the respective category using negative binomial models. We modelled the relative frequency, which considers the number of purchase occasions including the respective category in relation to overall purchasing frequency, by including an offset, i.e., log terms with a coefficient of 1, of overall purchasing frequency. Interaction terms between sociodemographic characteristics were explored but not included in final models as they were not statistically significant.

All data preparation and analysis tasks were carried out using R version 4.5.0. Alpha was set at 0.05.

### Deviations from the analysis plan

The analysis followed a previously published analysis plan, available here: https://osf.io/qfuvr/. A deviation from the protocol was to omit the analysis of purchasing frequency by region of residence, as we did not assess geographical variation in menus. We decided against adding more time points between June 2022 and October 2023 and included June data only to rule out seasonal differences in menus.

## Results

Tables 2 and 3 show the unadjusted and adjusted estimates of changes in calorie content and prevalence of lower-calorie drinks and mains under 600 kcal, respectively. Model coefficients of changes in calorie content are provided in Supplementary Material 2, forest plots overall and stratified by subgroup in Supplementary Material 3, and findings from the purchasing analysis in Supplementary Material 4.

**TABLE 2.**
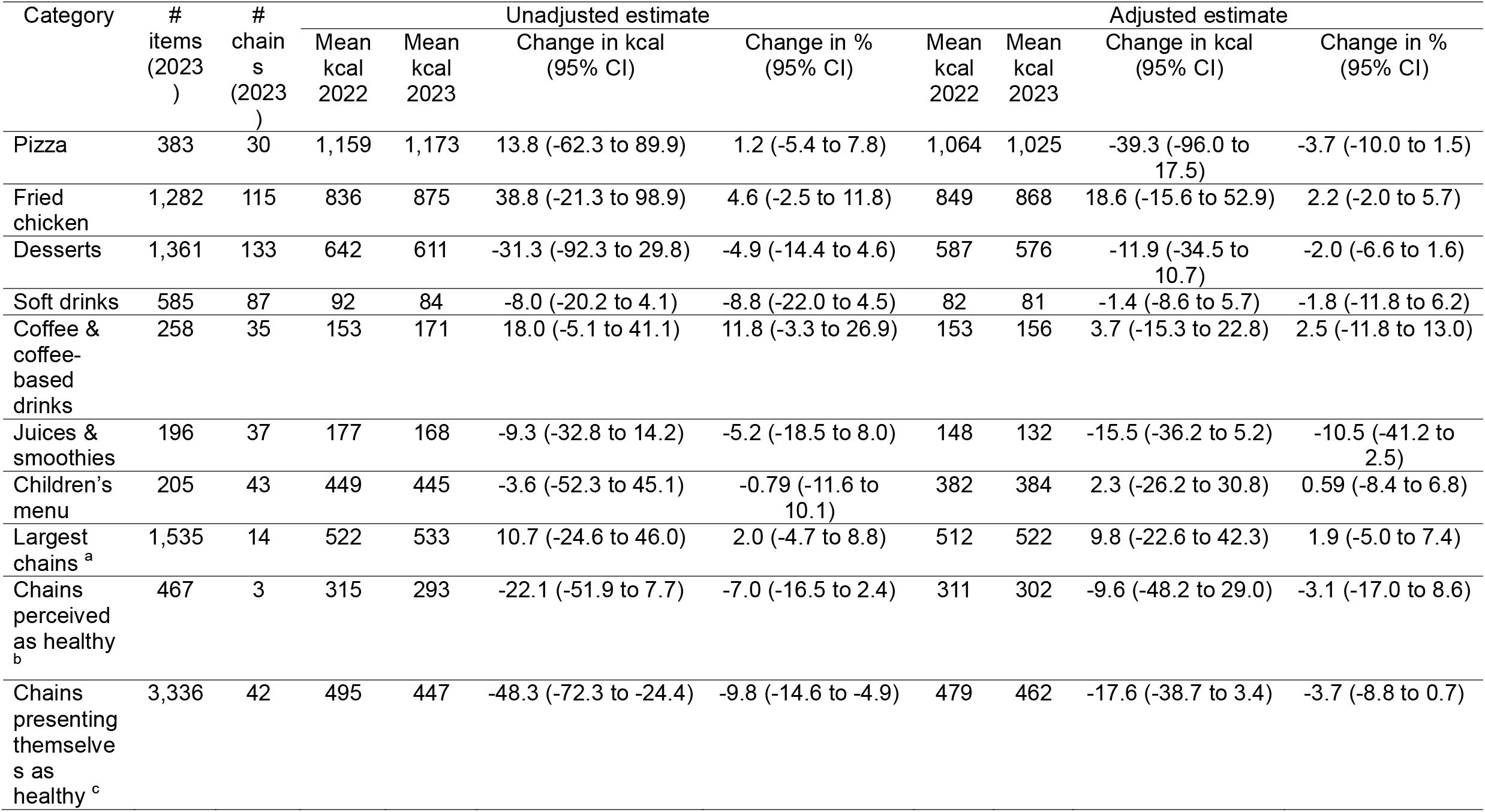
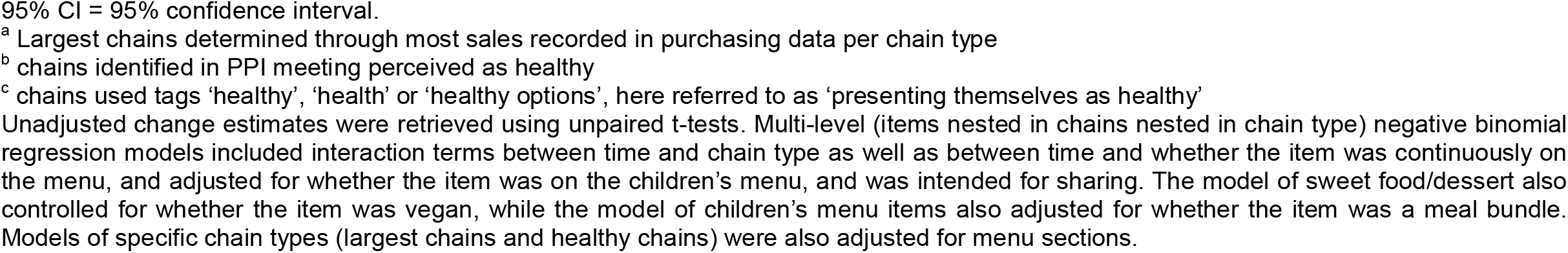
CHANGE IN CALORIE (KCAL) CONTENT BETWEEN JUNE 2022 AND JUNE 2023.

| Category | # items (2023) | # chains (2023) | Unadjusted estimate |  |  |  | Adjusted estimate |  |  |  |
| --- | --- | --- | --- | --- | --- | --- | --- | --- | --- | --- |
|  |  |  | Mean kcal 2022 | Mean kcal 2023 | Change in kcal (95% CI) | Change in % (95% CI) | Mean kcal 2022 | Mean kcal 2023 | Change in kcal (95% CI) | Change in % (95% CI) |
| Pizza | 383 | 30 | 1,159 | 1,173 | 13.8 (-62.3 to 89.9) | 1.2 (-5.4 to 7.8) | 1,064 | 1,025 | -39.3 (-96.0 to 17.5) | -3.7 (-10.0 to 1.5) |
| Fried chicken | 1,282 | 115 | 836 | 875 | 38.8 (-21.3 to 98.9) | 4.6 (-2.5 to 11.8) | 849 | 868 | 18.6 (-15.6 to 52.9) | 2.2 (-2.0 to 5.7) |
| Desserts | 1,361 | 133 | 642 | 611 | -31.3 (-92.3 to 29.8) | -4.9 (-14.4 to 4.6) | 587 | 576 | -11.9 (-34.5 to 10.7) | -2.0 (-6.6 to 1.6) |
| Soft drinks | 585 | 87 | 92 | 84 | -8.0 (-20.2 to 4.1) | -8.8 (-22.0 to 4.5) | 82 | 81 | -1.4 (-8.6 to 5.7) | -1.8 (-11.8 to 6.2) |
| Coffee & coffee-based drinks | 258 | 35 | 153 | 171 | 18.0 (-5.1 to 41.1) | 11.8 (-3.3 to 26.9) | 153 | 156 | 3.7 (-15.3 to 22.8) | 2.5 (-11.8 to 13.0) |
| Juices & smoothies | 196 | 37 | 177 | 168 | -9.3 (-32.8 to 14.2) | -5.2 (-18.5 to 8.0) | 148 | 132 | -15.5 (-36.2 to 5.2) | -10.5 (-41.2 to 2.5) |
| Children's menu | 205 | 43 | 449 | 445 | -3.6 (-52.3 to 45.1) | -0.79 (-11.6 to 10.1) | 382 | 384 | 2.3 (-26.2 to 30.8) | 0.59 (-8.4 to 6.8) |
| Largest chains <sup>a</sup> | 1,535 | 14 | 522 | 533 | 10.7 (-24.6 to 46.0) | 2.0 (-4.7 to 8.8) | 512 | 522 | 9.8 (-22.6 to 42.3) | 1.9 (-5.0 to 7.4) |
| Chains perceived as healthy <sup>b</sup> | 467 | 3 | 315 | 293 | -22.1 (-51.9 to 7.7) | -7.0 (-16.5 to 2.4) | 311 | 302 | -9.6 (-48.2 to 29.0) | -3.1 (-17.0 to 8.6) |
| Chains presenting themselves as healthy <sup>c</sup> | 3,336 | 42 | 495 | 447 | -48.3 (-72.3 to -24.4) | -9.8 (-14.6 to -4.9) | 479 | 462 | -17.6 (-38.7 to 3.4) | -3.7 (-8.8 to 0.7) |

95% CI = 95% confidence interval.
<sup>a</sup> Largest chains determined through most sales recorded in purchasing data per chain type
<sup>b</sup> chains identified in PPI meeting perceived as healthy
<sup>c</sup> chains used tags 'healthy', 'health' or 'healthy options', here referred to as 'presenting themselves as healthy'
Unadjusted change estimates were retrieved using unpaired t-tests. Multi-level (items nested in chains nested in chain type) negative binomial regression models included interaction terms between time and chain type as well as between time and whether the item was continuously on the menu, and adjusted for whether the item was on the children's menu, and was intended for sharing. The model of sweet food/dessert also controlled for whether the item was vegan, while the model of children's menu items also adjusted for whether the item was a meal bundle. Models of specific chain types (largest chains and healthy chains) were also adjusted for menu sections.

**TABLE 3.**
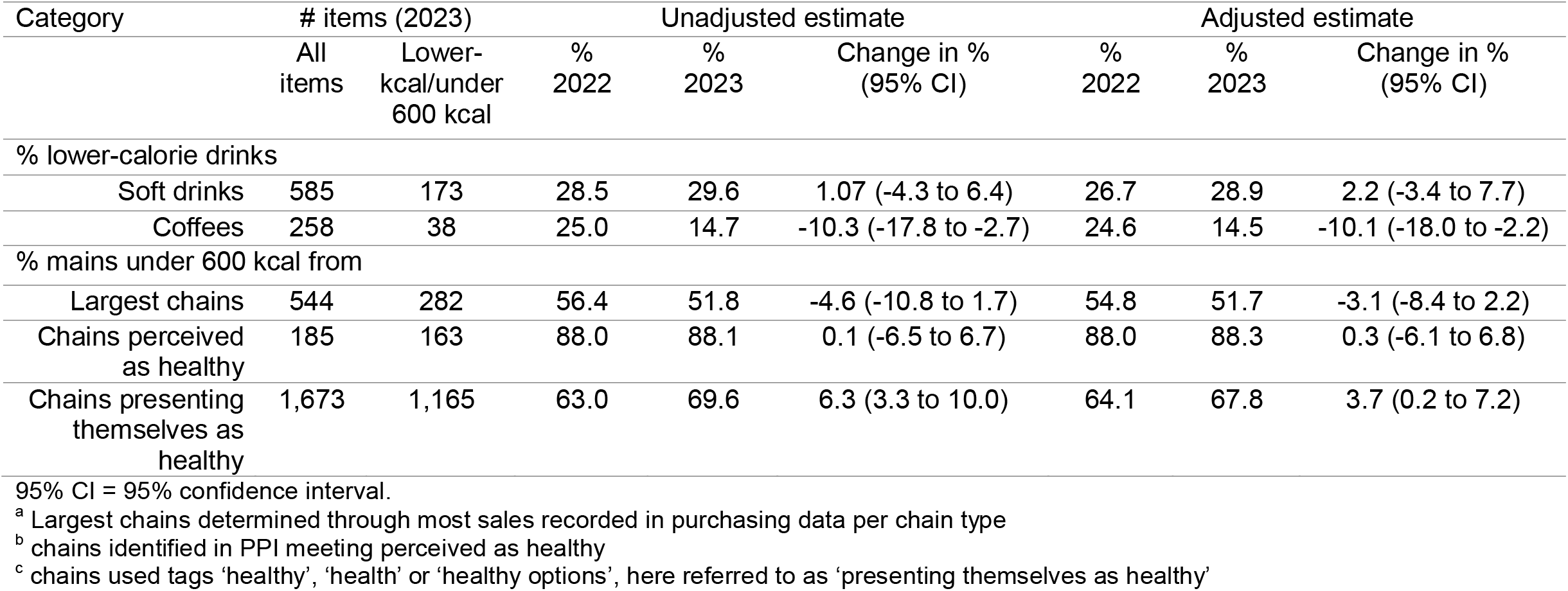
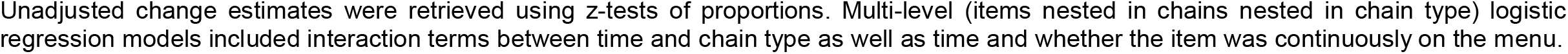
CHANGE IN PREVALENCE OF LOWER-CALORIE DRINKS AND MAINS UNDER 600 KCAL BETWEEN JUNE 2022 AND JUNE 2023.

| Category | # items (2023) |  | Unadjusted estimate |  |  | Adjusted estimate |  |  |
| --- | --- | --- | --- | --- | --- | --- | --- | --- |
|  | All items | Lower-kcal/under 600 kcal | % 2022 | % 2023 | Change in % (95% CI) | % 2022 | % 2023 | Change in % (95% CI) |
| % lower-calorie drinks |  |  |  |  |  |  |  |  |
| Soft drinks | 585 | 173 | 28.5 | 29.6 | 1.07 (-4.3 to 6.4) | 26.7 | 28.9 | 2.2 (-3.4 to 7.7) |
| Coffees | 258 | 38 | 25.0 | 14.7 | -10.3 (-17.8 to -2.7) | 24.6 | 14.5 | -10.1 (-18.0 to -2.2) |
| % mains under 600 kcal from |  |  |  |  |  |  |  |  |
| Largest chains | 544 | 282 | 56.4 | 51.8 | -4.6 (-10.8 to 1.7) | 54.8 | 51.7 | -3.1 (-8.4 to 2.2) |
| Chains perceived as healthy | 185 | 163 | 88.0 | 88.1 | 0.1 (-6.5 to 6.7) | 88.0 | 88.3 | 0.3 (-6.1 to 6.8) |
| Chains presenting themselves as healthy | 1,673 | 1,165 | 63.0 | 69.6 | 6.3 (3.3 to 10.0) | 64.1 | 67.8 | 3.7 (0.2 to 7.2) |

95% CI = 95% confidence interval.
<sup>a</sup> Largest chains determined through most sales recorded in purchasing data per chain type
<sup>b</sup> chains identified in PPI meeting perceived as healthy
<sup>c</sup> chains used tags 'healthy', 'health' or 'healthy options', here referred to as 'presenting themselves as healthy'
Unadjusted change estimates were retrieved using z-tests of proportions. Multi-level (items nested in chains nested in chain type) logistic regression models included interaction terms between time and chain type as well as time and whether the item was continuously on the menu.

Among chains presenting themselves as healthy, average calories per item decreased from 495 kcal/item in June 2022 to 447 kcal/item in June 2023, while the adjusted estimate was attenuated (-17.6 kcal/item, 95% CI -38.7 to 3.4). In addition, these chains increased the proportion of main meals under 600 kcal from 63.0% to 69.6% with an adjusted difference of 3.7 percentage points (95% CI 0.2 to 7.2). This increase was predominantly driven by Fast-Food and Takeaway chains and by items not continuously on the menu (see Figure 3). A higher frequency of purchasing from chains presenting themselves as healthy was observed for men (OR 1.19, 95%CI 1.02 to 1.40), people aged 35–44 years (e.g. >64 years OR 0.67, 95% CI 0.52 to 0.86 compared to 35–44 years), and individuals with higher SES (low SES OR 0.63, 95%CI 0.49 to 0.81 compared to high SES). Trends in relative purchasing were similar for age and SES, but there was no difference in relative purchasing frequency by sex.

**Figure 3.**
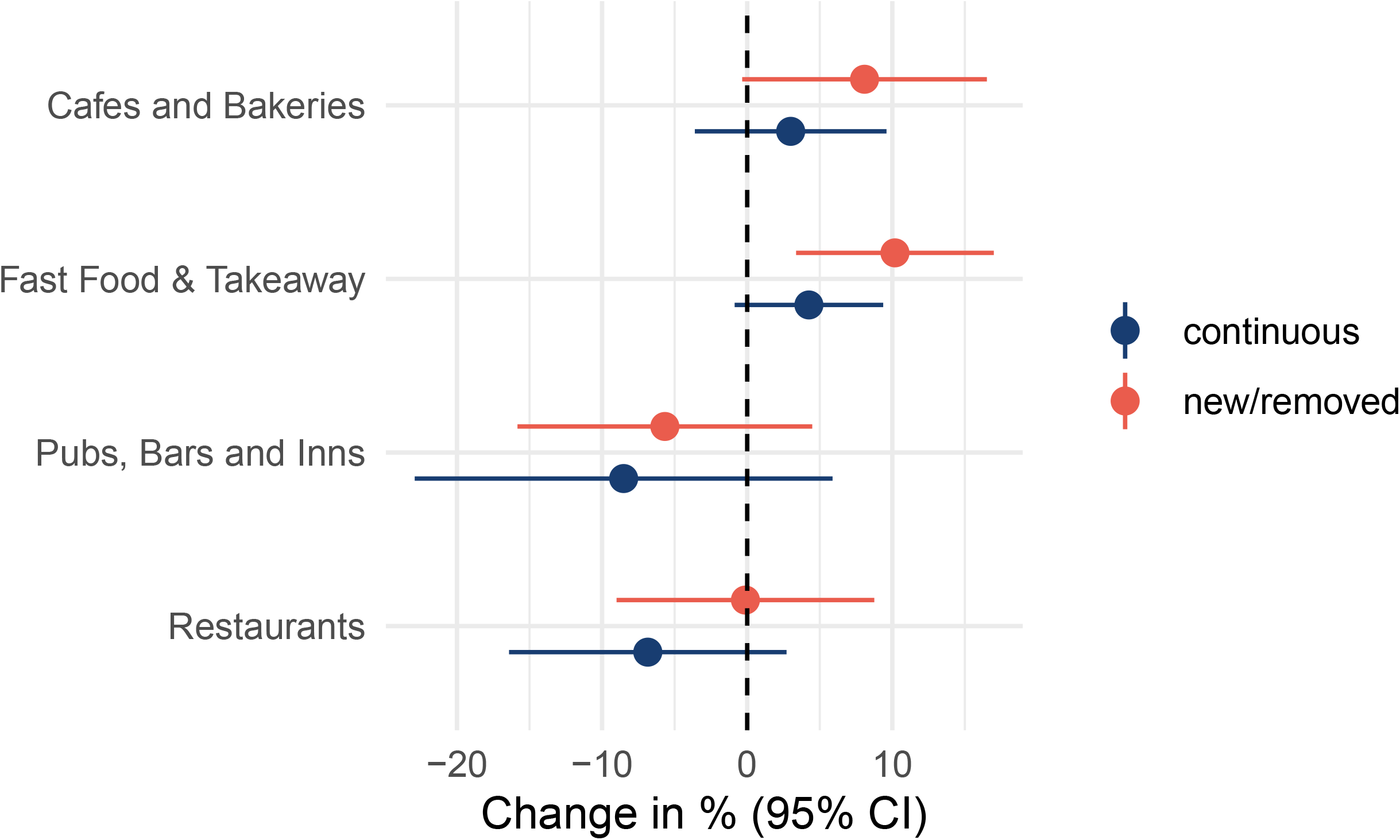
Change in the percentage of main food items under 600 kcal from chains presenting themselves as healthy between June 2022 and June 2023. Estimates were retrieved from a multilevel logistic model including interaction terms between time and chain type and whether the item was continuously on the menu.

The share of lower-calorie coffees decreased by 10 pp (95% CI -18.0 to -0.02). The frequency of purchasing coffee was higher among men compared to women (OR 1.30, 95%CI 1.14 to 1.48) and increased with age (e.g. over 64 years OR 2.13, 95% CI 1.74 to 2.61 compared to 35–44 years).

We found no overall change in any of the other investigated categories, but change was observed in some subgroups (see Supplementary Material 3). New pizzas in pubs (only in 2023) contained 226 kcal less (95% CI -442.0 to -10.1) on average compared to removed pizzas (only in 2022). Absolute purchasing frequency was 91% higher for men (95% CI 1.09 to 3.43) and increased with age, while there was no difference in relative purchasing.

We observed a mean reduction of 43.6 kcal (95% CI -75.8 to -11.5) per sweet food/dessert among Fast-Food & Takeaway chains. Purchasing decreased with age, while relative purchasing was lower among men (OR 0.82, 95% CI 0.73 to 0.93).

New juices and smoothies contained on average 41 kcal less (95% CI -76.1 to -5.2) than removed ones. Increasing age was associated with higher absolute purchasing of these products (e.g. 65+ years OR 1.60, 95% CI 1.10 to 2.35), but with lower relative purchasing (e.g. 55–64 years: OR 0.59, 95% CI 0.41 to 0.84 compared to 35–44 years).

Among the largest chains, the calorie content of drinks increased by 22 kcal (95% CI 3.1 to 41) on average. Increasing age was associated with higher absolute purchasing but lower relative purchasing, while absolute purchasing was on average 13% higher for men (95% CI 1.04 to 1.23).

### Sensitivity analysis

Supplementary Material 5 contains results from the analysis excluding items for which multiple calorie values were present in the menu data. Findings were mostly robust, with a notable exception of pizzas purchased from pubs, with the formerly observed calorie reduction no longer statistically significant. Similarly, findings for chains presenting themselves as healthy were similar to the main analysis, but the increase in mains under 600 kcal was no longer statistically significant in the adjusted analysis except among Fast-Food and Takeaway chains.

## Discussion

### Summary of findings

This study investigated menu changes during one year after the implementation of calorie labelling and who may be most affected by these changes in Great Britain. Among the investigated categories, we observed limited change in the calorie content of menu items over time. We found some evidence of structural changes, with a decrease in the proportion of lower-calorie coffees, which may affect men and older people as the most frequent coffee purchasers. We also observed an increase in the proportion of main food items under 600 kcal among chains that present themselves as healthy, with men, people aged between 35 and 44 years and with high SES, who purchased from these chains most frequently, potentially benefitting most. Findings were partially sensitive to methodological choices, likely due to reduced statistical power resulting from smaller samples in the sensitivity analysis. Findings relating to purchasing need to be interpreted with caution, as only 15 out of the 43 chains could be identified in the purchase data.

### Comparison with previous literature

Despite the mixed evidence on overall calorie reduction [15,23,24], research generally reports that changes in calories tend to be driven by menu changes such as the removal of higher-calorie dishes [16,25], as well as the introduction of lower-calorie dishes [26–28], rather than reformulating items that are continuously on the menu [29]. Our findings corroborate these observations.

While the majority of previous research reports on varying menu changes by the type of item, the evidence base remains largely mixed [16,30]. For instance, our findings align with previous research reporting no calorie changes among pizzas and children’s items from the US context [15,30], while the evidence on calorie changes in coffee is mixed [31,32]. A previous meta-analysis of US studies [15] did not find different calorie reductions among ‘healthier’ menu items, but their item-level classification was different from our definition which relied on tags assigned by chains themselves.

The international evidence on whether menu changes following calorie disclosure depend on the type of food outlet is mixed [23,25,33]. The only study to date from the English context reports a mean reduction of 38 kcal (95% CI -42 to -5) per menu item for sit-down restaurants, and a 42 kcal/item (95% CI 27 to 57) increase among fast-food outlets [16]. In contrast, we did not observe differences in the change in calorie content by chain type among most of the categories examined, apart from greater calorie reductions among Fast-Food and Takeaway chains presenting themselves as healthy. This may be due to systematic differences between chains in general and those marketing themselves as healthy. Our analysis of differences in purchasing of the selected categories is an important contribution in gauging the potential differential impacts of menu changes. Our investigation of both absolute and relative purchasing frequency allows to explore potential population-wide impact while accounting for higher overall purchasing frequency by some population groups.

### Limitations

Despite using comprehensive online menu data and consumer purchase data from Great Britain, several limitations apply. By design, no pre-policy data were available, as platforms only began posting calorie information at a large scale when the calorie labelling regulations were in place. Consequently, we could capture the impact of neither the policy’s implementation nor anticipatory effects on chains’ menus, but, instead, we followed trends after policy implementation. Our exploratory analysis, without a control group, cannot establish causality, but serve as starting point for further research. Limited data availability over time means that we were only able to analyse menu changes over one year, while accounting for seasonal variation. It is plausible that menu changes are a continuous, slow process which may have been too small over one year to be captured in our analysis. In addition, the accuracy of calorie labelling on online platforms over time is unknown, and no information was available on portion sizes, precluding analyses of whether calorie reductions were due to reducing portion sizes or reformulating dishes.

### Interpretation of findings

Our findings contribute to a growing evidence base that England’s calorie labelling regulations may have a small but positive impact in reducing calories on menus [16,27]. From a theoretical perspective, a structural change that does not require individual behaviour change is more successful in reducing population calorie intake compared to an intervention requiring consumers to notice and act on nutritional information [34,35]. Although menu changes in the present study were limited, likely due to the lack of pre-implementation data, previous UK research found a mean reduction of 9 kcal (95% CI -16 to -1) per item overall [16]. Taken together, the evidence suggests the occurrence of menu changes following mandatory calorie labelling, presenting an opportunity for structural change in the food environment.

A potential reason for the small extent of observed menu changes is that the regulations require calories to be shown, not reduced [6]. Previous voluntary efforts which entailed both disclosure and reduction of calories achieved limited success [10]. It is plausible that public health concerns are less relevant to business practices than economic targets, customer satisfaction, and reputation. For instance, qualitative findings from a large-scale evaluation found that businesses in England tended to be reluctant to introduce noticeable menu changes and preferred ‘health-by-stealth’ approaches [36]. This may also explain why the only overall calorie reduction in our study was observed among chains presenting themselves as healthy, as such businesses may specifically target customers interested in healthy, and potentially lower-calorie foods and drinks.

To the best of our knowledge, this was the first study to explore how menu changes may affect population health by considering purchasing patterns. For chains presenting themselves as healthy, associations with consumer characteristics need to be interpreted with caution as we could identify only 35% respective chains in the purchase data. However, in addition to our finding of individuals with higher SES purchasing from these chains more frequently, it is plausible that consumers interested in healthy food already have healthier diets than consumers who are not [37]. Thereby, this reduction would at best not alter dietary health inequalities, and at worst widen them.

### Implications for research and policy

Future research should continue to monitor calories and other nutritional indicators in the OOH food sector in the long term, including the accuracy of reporting. Such an objective links well with the recently published 10-year Health Plan in the UK, which champions healthy food sales monitoring [38]. Given the greater success of structural interventions on population health [34], research may ascertain the effectiveness of calorie reduction efforts targeted at menu items on offer such as fiscal incentives encouraging lower calorie content relative to portion size, akin to the Soft Drinks Industry Levy [39].

Finally, while calorie labelling may be viewed as beneficial through information provision irrespective of its impact on calories consumed, it is essential to understand the harms it may cause, particularly to people with experience of disordered eating and eating disorders [40]. Future research as well as stakeholders tasked with the continuation, review, and adaptation need to consider the policy’s observed benefits and harms jointly.

## Conclusions

Our findings contribute to a growing evidence base on calorie labelling in the OOH food sector. On the one hand, menu change may have the potential for small improvements in population diet and does not rely on individual behaviour change. On the other hand, this study’s findings suggest that if menu changes are concentrated among businesses offering ‘healthier’, already lower-calorie food, only, diet-related health inequalities may widen.

## Data Availability

Web-scraping tools used to collect online menu data are publicly available. Code is available upon request. Data are available upon request subject to a data sharing agreement. Use of these data is only permitted for non-commercial purposes. Worldpanel by Numerator, GB Out of Home Panel data used in this study cannot be shared but can be obtained from Worldpanel by Numerator (https://worldpanelbynumerator.com). Data were available for 1st January to 31st December 2022. The analyses used data from 3rd January to 27th November 2022. All analyses and interpretation were conducted independently of Worldpanel by Numerator.

