## Supplementary Material 1 for "How have restaurant menus changed following England’s calorie labelling regulations and who is likely to benefit? A longitudinal analysis of online menu data"

### Supplementary Material 1: Public and policy involvement in the CARO project

Throughout this project (https://nihrsphr.link/CARO), researchers worked closely with two involvement/advisory panels, specifically a public and policy involvement panel. Public involvement in research can be defined as research being carried out 'with' or 'by' members of the public rather than 'to', 'about' or 'for' them [1]. The benefits of public and policy involvement are well documented [2], from the research conceptualisation to dissemination stage [3]. In this project, we selected relevant food groups and chains to prioritise jointly with our public and policy advisors. Prioritising categories in this way enabled our research to be highly relevant and impactful to the public and policymakers in this area.

Our public involvement panel consisted of six members of the public with experience in providing advice on studies relating to food policy and sustainable diets, recruited through the Dietary Change for Greater Environmental Sustainability (DIGEST) Public Involvement Panel based at the London School of Hygiene & Tropical Medicine. The policy panel consisted of seven national and local-level public health professionals with a remit in food and obesity policy, recruited through public health networks.

Figure 1 depicts the panels' involvement throughout the stages of the project. Both public and policy involvement panels provided input into the category selection during online workshops. These workshops were structured along the prompts shown in Figure 1. In addition, panels were shown findings from previous research and asked to relate the workshop's findings to these. At the end of each workshop, we asked panellists to prioritise among the possible categories. We included categories deemed most relevant by each panel and the literature, prioritising those raised by more than one of these three, and finally excluding those that could not be realised using the available data. With respect to the latter, we excluded the categories suggested by the public advisors 'types of protein' and 'cooking methods of protein' as these could not be determined from the items' names and descriptions in the available menu data.

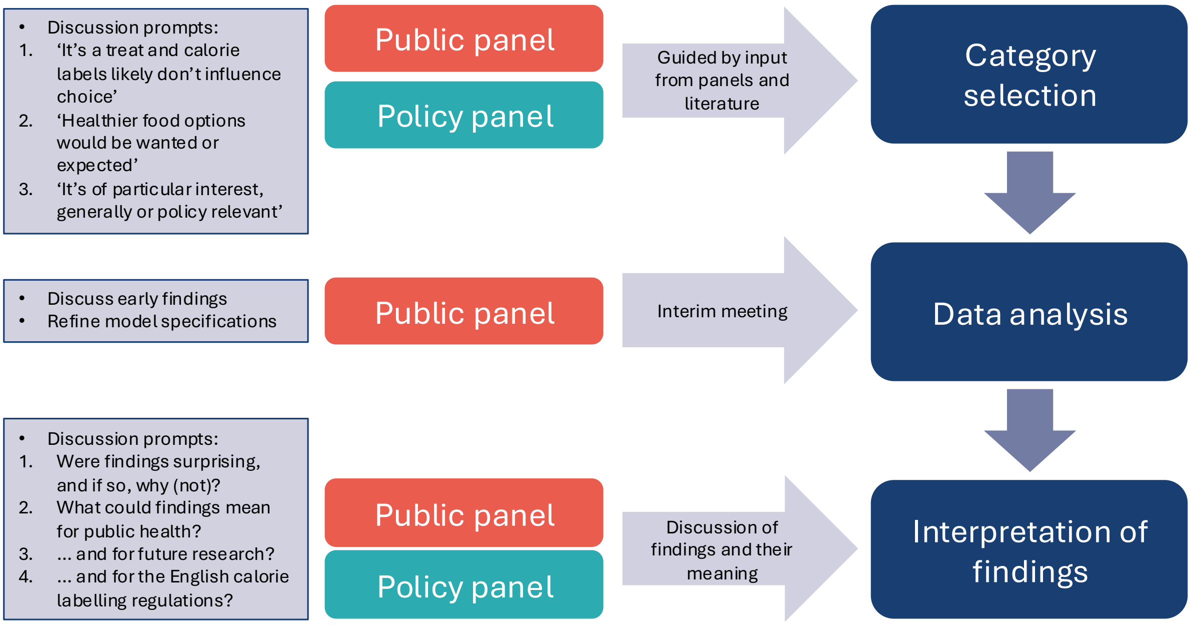


**Figure 1.** Study overview.

During the analysis stage, researchers met again with the public panel to discuss early findings and refine the analysis as needed. As a result of this meeting, the drink category was split further into soft drinks, coffees, and juices/smoothies, as opposed to cold and hot beverages. Another change that emerged from this meeting was to split sweet foods/deserts further into vegan and non-vegan items as the latter may be (advertised as) healthier and therefore lower in calorie content.

After the analysis, researchers met again with both panels in online workshops to discuss the findings and their implications. Specifically, panellists were asked 1) if findings were surprising and why (not); 2) what findings could mean for public health, 3) what future research could be done, and 4) what are the implications for the calorie labelling regulations in England? The final workshop with the public panel was attended by a visual scribe who captured key points raised during the meeting in a visual summary (https://nihrsphr.link/CARO).
