## Supplementary Material 2 for "How have restaurant menus changed following England’s calorie labelling regulations and who is likely to benefit? A longitudinal analysis of online menu data"

### Supplementary Material 2: Model coefficients from multi-level models predicting the change in calorie content

**Pizza**

| | kcal | | |
|---|---|---|---|
| *Predictors* | *Incidence Rate Ratios* | *CI* | *p* |
| (Intercept) | 430.25 | 309.39 – 598.33 | **<0.001** |
| year [2023] | 1.13 | 0.77 – 1.67 | 0.527 |
| chain type [Fast Food & Takeaway] | 2.37 | 1.60 – 3.53 | **<0.001** |
| chain type [Pizza chain] | 2.33 | 1.63 – 3.33 | **<0.001** |
| chain type [Pubs, Bars and Inns] | 2.15 | 1.41 – 3.28 | **<0.001** |
| chain type [Restaurants] | 2.57 | 1.78 – 3.72 | **<0.001** |
| continuous [new/removed] | 1.06 | 0.98 – 1.15 | 0.157 |
| children | 0.48 | 0.38 – 0.61 | **<0.001** |
| sharing bundle | 1.29 | 1.15 – 1.45 | **<0.001** |
| year [2023] × chain type [Fast Food & Takeaway] | 0.94 | 0.61 – 1.44 | 0.773 |
| year [2023] × chain type [Pizza chain] | 0.85 | 0.58 – 1.26 | 0.430 |
| year [2023] × chain type [Pubs, Bars and Inns] | 0.70 | 0.45 – 1.10 | 0.120 |
| year [2023] × chain type [Restaurants] | 0.89 | 0.60 – 1.34 | 0.587 |
| year [2023] × continuous [new/removed] | 0.97 | 0.87 – 1.09 | 0.593 |
| **Random Effects** | | | |
| $\sigma^2$ | | | |
| $\tau_{00\ chain:chain\_type}$ | 0.05 | | |
| $\tau_{00\ chain\_type}$ | 0.00 | | |
| $N_{chain}$ | 33 | | |
| $N_{chain\_type}$ | 5 | | |
| Observations | 805 | | |

**Fried chicken**

| | kcal | | |
|---|---|---|---|
| *Predictors* | *Incidence Rate Ratios* | *CI* | *p* |
| (Intercept) | 697.58 | 499.00 – 975.20 | **<0.001** |
| year [2023] | 1.08 | 0.81 – 1.44 | 0.601 |
| chain type [Chicken chain] | 1.27 | 0.88 – 1.83 | 0.207 |
| chain type [Fast Food & Takeaway] | 1.20 | 0.85 – 1.70 | 0.310 |
| chain type [Pubs, Bars and Inns] | 1.75 | 1.17 – 2.62 | **0.006** |
| chain type [Restaurants] | 1.29 | 0.90 – 1.84 | 0.172 |
| continuous [new/removed] | 1.06 | 0.99 – 1.13 | 0.103 |
| children | 0.54 | 0.49 – 0.60 | **<0.001** |
| sharing bundle [Fried chicken only for 1] | 0.68 | 0.64 – 0.71 | **<0.001** |
| sharing bundle [Sharing dish] | 2.59 | 2.39 – 2.80 | **<0.001** |
| year [2023] × chain type [Chicken chain] | 0.94 | 0.70 – 1.26 | 0.684 |
| year [2023] × chain type [Fast Food & Takeaway] | 0.91 | 0.67 – 1.22 | 0.511 |
| year [2023] × chain type [Pubs, Bars and Inns] | 0.99 | 0.71 – 1.38 | 0.953 |
| year [2023] × chain type [Restaurants] | 0.93 | 0.68 – 1.27 | 0.659 |
| year [2023] × continuous [new/removed] | 1.04 | 0.95 – 1.13 | 0.412 |
| **Random Effects** | | | |
| $\sigma^2$ | | | |
| $\tau_{00\ chain:chain\_type}$ | 0.11 | | |
| $\tau_{00\ chain\_type}$ | 0.00 | | |
| $N_{chain}$ | 123 | | |
| $N_{chain\_type}$ | 5 | | |
| Observations | 2446 | | |

**Sweet food and desserts**

| | kcal | | |
|---|---|---|---|
| *Predictors* | *Incidence Rate Ratios* | *CI* | *p* |
| (Intercept) | 390.14 | 329.10 – 462.50 | **<0.001** |
| year [2023] | 1.00 | 0.93 – 1.08 | 0.928 |
| chain type [Fast Food & Takeaway] | 1.07 | 0.88 – 1.30 | 0.509 |
| chain type [Pubs, Bars and Inns] | 2.28 | 1.61 – 3.22 | **<0.001** |
| chain type [Restaurants] | 1.40 | 1.12 – 1.74 | **0.003** |
| chain type [Sweet food/ Dessert Shop] | 1.95 | 1.45 – 2.63 | **<0.001** |
| continuous [new/removed] | 1.06 | 1.00 – 1.12 | 0.059 |
| vegan sweet dessert | 0.79 | 0.72 – 0.87 | **<0.001** |
| sharing | 3.18 | 2.84 – 3.57 | **<0.001** |
| year [2023] × chain type [Fast Food & Takeaway] | 0.90 | 0.82 – 1.00 | **0.042** |
| year [2023] × chain type [Pubs, Bars and Inns] | 0.94 | 0.74 – 1.19 | 0.611 |
| year [2023] × chain type [Restaurants] | 1.05 | 0.92 – 1.19 | 0.463 |
| year [2023] × chain type [Sweet food/ Dessert Shop] | 1.00 | 0.91 – 1.10 | 0.965 |
| year [2023] × continuous [new/removed] | 0.98 | 0.91 – 1.06 | 0.597 |
| year [2023] × vegan sweet dessert | 1.08 | 0.95 – 1.23 | 0.228 |
| **Random Effects** | | | |
| $\sigma^2$ | | | |
| $\tau_{00\ chain:chain\_type}$ | 0.13 | | |
| $\tau_{00\ chain\_type}$ | 0.00 | | |
| $N_{chain}$ | 140 | | |
| $N_{chain\_type}$ | 5 | | |
| Observations | 2656 | | |

**Soft drinks (change in kcal)**

| | kcal | | |
|---|---|---|---|
| *Predictors* | *Incidence Rate Ratios* | *CI* | *p* |
| (Intercept) | 111.19 | 85.18 – 145.14 | **<0.001** |
| year [2023] | 1.05 | 0.88 – 1.26 | 0.588 |
| chain type [Fast Food & Takeaway] | 0.95 | 0.70 – 1.29 | 0.747 |
| chain type [Pubs, Bars and Inns] | 0.75 | 0.37 – 1.50 | 0.411 |
| chain type [Restaurants] | 1.04 | 0.74 – 1.46 | 0.817 |
| continuous [new/removed] | 1.12 | 0.97 – 1.29 | 0.134 |
| sugar free soft drink | 0.02 | 0.02 – 0.02 | **<0.001** |
| year [2023] × chain type [Fast Food & Takeaway] | 0.97 | 0.79 – 1.20 | 0.784 |
| year [2023] × chain type [Pubs, Bars and Inns] | 0.99 | 0.52 – 1.90 | 0.978 |
| year [2023] × chain type [Restaurants] | 0.94 | 0.73 – 1.21 | 0.633 |
| year [2023] × continuous [new/removed] | 0.92 | 0.77 – 1.10 | 0.355 |
| **Random Effects** | | | |
| $\sigma^2$ | | | |
| $\tau_{00\ chain:chain\_type}$ | 0.20 | | |
| $\tau_{00\ chain\_type}$ | 0.00 | | |
| $N_{chain}$ | 98 | | |
| $N_{chain\_type}$ | 4 | | |
| Observations | 1171 | | |

**Soft drinks (prevalence of low-sugar/sugar-free)**

| | sugar_free_soft_drink | | |
|---|---|---|---|
| *Predictors* | *Odds Ratios* | *CI* | *p* |
| (Intercept) | 0.26 | 0.14 – 0.48 | **<0.001** |
| year [2023] | 1.07 | 0.56 – 2.05 | 0.835 |
| chain type [Fast Food & Takeaway] | 1.90 | 0.94 – 3.81 | 0.072 |
| chain type [Pubs, Bars and Inns] | 0.93 | 0.14 – 6.31 | 0.941 |
| chain type [Restaurants] | 1.21 | 0.54 – 2.72 | 0.638 |
| continuous [new/removed] | 0.91 | 0.58 – 1.41 | 0.658 |
| year [2023] × chain type [Fast Food & Takeaway] | 0.91 | 0.44 – 1.86 | 0.793 |
| year [2023] × chain type [Pubs, Bars and Inns] | 1.11 | 0.13 – 9.67 | 0.928 |
| year [2023] × chain type [Restaurants] | 1.02 | 0.43 – 2.44 | 0.956 |
| year [2023] × continuous [new/removed] | 1.22 | 0.69 – 2.15 | 0.487 |
| **Random Effects** | | | |
| $\sigma^2$ | 3.29 | | |
| $\tau_{00\ chain:chain\_type}$ | 0.52 | | |
| $\tau_{00\ chain\_type}$ | 0.00 | | |
| $N_{chain}$ | 98 | | |
| $N_{chain\_type}$ | 4 | | |
| Observations | 1171 | | |
| Marginal $R^2$ / Conditional $R^2$ | 0.021 / NA | | |

**Coffee and coffee-based drinks (change in kcal)**

| | kcal | | |
|---|---|---|---|
| *Predictors* | *Incidence Rate Ratios* | *CI* | *p* |
| (Intercept) | 185.49 | 146.66 – 234.60 | **<0.001** |
| year [2023] | 1.07 | 0.86 – 1.34 | 0.523 |
| chain type [Coffee Shop] | 1.14 | 0.81 – 1.61 | 0.440 |
| chain type [Fast Food & Takeaway] | 0.85 | 0.61 – 1.17 | 0.317 |
| chain type [Restaurants] | 0.43 | 0.25 – 0.73 | **0.002** |
| continuous [new/removed] | 1.07 | 0.88 – 1.30 | 0.510 |
| lower kcal coffee | 0.04 | 0.04 – 0.05 | **<0.001** |
| year [2023] × chain type [Coffee Shop] | 0.83 | 0.63 – 1.10 | 0.187 |
| year [2023] × chain type [Fast Food & Takeaway] | 0.95 | 0.70 – 1.27 | 0.715 |
| year [2023] × chain type [Restaurants] | 1.21 | 0.74 – 1.97 | 0.442 |
| year [2023] × continuous [new/removed] | 1.07 | 0.84 – 1.37 | 0.566 |
| **Random Effects** | | | |
| $\sigma^2$ | | | |
| $\tau_{00\ chain:chain\_type}$ | 0.06 | | |
| $\tau_{00\ chain\_type}$ | 0.00 | | |
| $N_{chain}$ | 39 | | |
| $N_{chain\_type}$ | 4 | | |
| Observations | 482 | | |

**Coffee and coffee-based drinks (prevalence of lower-calorie coffees)**

| | lower_kcal_coffee | | |
|---|---|---|---|
| *Predictors* | *Odds Ratios* | *CI* | *p* |
| (Intercept) | 0.26 | 0.11 – 0.59 | **0.001** |
| year [2023] | 0.99 | 0.37 – 2.62 | 0.981 |
| chain type [Coffee Shop] | 1.23 | 0.42 – 3.63 | 0.701 |
| chain type [Fast Food & Takeaway] | 0.90 | 0.29 – 2.81 | 0.858 |
| chain type [Restaurants] | 6.22 | 1.25 – 30.88 | **0.025** |
| continuous [new/removed] | 0.96 | 0.44 – 2.11 | 0.922 |
| year [2023] × chain type [Coffee Shop] | 0.99 | 0.28 – 3.47 | 0.982 |
| year [2023] × chain type [Fast Food & Takeaway] | 1.24 | 0.32 – 4.78 | 0.756 |
| year [2023] × chain type [Restaurants] | 0.05 | 0.00 – 0.64 | **0.021** |
| year [2023] × continuous [new/removed] | 0.28 | 0.09 – 0.88 | **0.028** |
| **Random Effects** | | | |
| $\sigma^2$ | 3.29 | | |
| $\tau_{00}$ chain:chain_type | 0.28 | | |
| $\tau_{00}$ chain_type | 0.00 | | |
| N chain | 39 | | |
| N chain_type | 4 | | |
| Observations | 482 | | |
| Marginal $R^2$ / Conditional $R^2$ | 0.157 / NA | | |

**Juice and smoothies**

| | **kcal** | | |
|---|---|---|---|
| *Predictors* | *Incidence Rate Ratios* | *CI* | *p* |
| (Intercept) | 109.21 | 68.43 – 174.31 | **<0.001** |
| year [2023] | 0.90 | 0.69 – 1.19 | 0.466 |
| chain type [Fast Food & Takeaway] | 1.27 | 0.67 – 2.44 | 0.463 |
| chain type [Juice Shop] | 1.71 | 0.63 – 4.64 | 0.294 |
| chain type [Restaurants] | 1.23 | 0.58 – 2.64 | 0.591 |
| continuous [new/removed] | 1.06 | 0.85 – 1.33 | 0.597 |
| year [2023] × chain type [Fast Food & Takeaway] | 1.07 | 0.71 – 1.61 | 0.756 |
| year [2023] × chain type [Juice Shop] | 1.14 | 0.80 – 1.62 | 0.469 |
| year [2023] × chain type [Restaurants] | 1.36 | 0.82 – 2.25 | 0.235 |
| year [2023] × continuous [new/removed] | 0.70 | 0.49 – 0.98 | **0.040** |
| **Random Effects** | | | |
| $\sigma^2$ | 0.39 | | |
| $\tau_{00\ chain:chain\_type}$ | 0.59 | | |
| $\tau_{00\ chain\_type}$ | 0.00 | | |
| ICC | 0.60 | | |
| $N_{chain}$ | 44 | | |
| $N_{chain\_type}$ | 4 | | |
| Observations | 428 | | |
| Marginal $R^2$ / Conditional $R^2$ | 0.068 / 0.632 | | |

**Children's menu**

| | kcal | | |
|---|---|---|---|
| *Predictors* | *Incidence Rate Ratios* | *CI* | *p* |
| (Intercept) | 306.02 | 177.88 – 526.47 | **<0.001** |
| year [2023] | 1.01 | 0.72 – 1.41 | 0.970 |
| chain type [Fast Food & Takeaway] | 1.32 | 0.73 – 2.39 | 0.358 |
| chain type [Pubs, Bars and Inns] | 1.18 | 0.56 – 2.51 | 0.666 |
| chain type [Restaurants] | 1.42 | 0.77 – 2.62 | 0.262 |
| continuous [new/removed] | 0.90 | 0.79 – 1.03 | 0.122 |
| drink | 0.23 | 0.16 – 0.33 | **<0.001** |
| meal bundle | 1.01 | 0.74 – 1.37 | 0.967 |
| year [2023] × chain type [Fast Food & Takeaway] | 1.03 | 0.73 – 1.45 | 0.877 |
| year [2023] × chain type [Pubs, Bars and Inns] | 1.01 | 0.68 – 1.49 | 0.968 |
| year [2023] × chain type [Restaurants] | 0.92 | 0.65 – 1.31 | 0.650 |
| year [2023] × continuous [new/removed] | 1.09 | 0.92 – 1.29 | 0.320 |
| year [2023] × meal bundle | 0.99 | 0.80 – 1.22 | 0.899 |
| **Random Effects** | | | |
| $\sigma^2$ | | | |
| $\tau_{00\ \text{chain:chain\_type}}$ | 0.33 | | |
| $\tau_{00\ \text{chain\_type}}$ | 0.00 | | |
| $N_{\text{chain}}$ | 49 | | |
| $N_{\text{chain\_type}}$ | 4 | | |
| Observations | 409 | | |

**Largest chains**

| | kcal | | |
|---|---|---|---|
| *Predictors* | *Incidence Rate Ratios* | *CI* | *p* |
| (Intercept) | 713.60 | 540.41 – 942.30 | **<0.001** |
| year [2023] | 1.03 | 0.84 – 1.27 | 0.756 |
| chain type [Fast Food & Takeaway] | 1.20 | 0.87 – 1.67 | 0.272 |
| chain type [Pubs, Bars and Inns] | 2.39 | 1.67 – 3.43 | **<0.001** |
| chain type [Restaurants] | 1.85 | 1.33 – 2.58 | **<0.001** |
| continuous [new/removed] | 0.94 | 0.86 – 1.03 | 0.200 |
| menu section [Children's Menu] | 0.17 | 0.13 – 0.23 | **<0.001** |
| menu section [Drinks] | 0.14 | 0.12 – 0.16 | **<0.001** |
| menu section [Mains] | 0.58 | 0.51 – 0.66 | **<0.001** |
| menu section [Starters/Sides] | 0.30 | 0.26 – 0.36 | **<0.001** |
| menu section [Sweets/Dessert] | 0.45 | 0.38 – 0.53 | **<0.001** |
| menu section [Toppings/Extras] | 0.26 | 0.19 – 0.36 | **<0.001** |
| year [2023] × chain type [Fast Food & Takeaway] | 0.93 | 0.79 – 1.10 | 0.396 |
| year [2023] × chain type [Pubs, Bars and Inns] | 0.89 | 0.71 – 1.11 | 0.291 |
| year [2023] × chain type [Restaurants] | 0.92 | 0.76 – 1.12 | 0.409 |
| year [2023] × continuous [new/removed] | 1.14 | 1.01 – 1.29 | **0.037** |
| year [2023] × menu section [Children's Menu] | 1.00 | 0.69 – 1.45 | 0.998 |
| year [2023] × menu section [Drinks] | 1.14 | 0.92 – 1.40 | 0.231 |
| year [2023] × menu section [Mains] | 1.03 | 0.87 – 1.22 | 0.727 |
| year [2023] × menu | 1.03 | 0.83 – 1.27 | 0.811 |

| | | | |
|---|---|---|---|
| section [Starters/Sides] | | | |
| year [2023] × menu section [Sweets/Dessert] | 0.95 | 0.77 – 1.18 | 0.660 |
| year [2023] × menu section [Toppings/Extras] | 0.82 | 0.55 – 1.24 | 0.354 |
| **Random Effects** | | | |
| $\sigma^2$ | | | |
| $\tau_{00\ chain:chain\_type}$ | 0.04 | | |
| $\tau_{00\ chain\_type}$ | 0.00 | | |
| $N_{chain}$ | 14 | | |
| $N_{chain\_type}$ | 4 | | |
| Observations | 2894 | | |

**Largest chains (prevalence of mains < 600 kcal)**

| | under600 | | |
|---|---|---|---|
| *Predictors* | *Odds Ratios* | *CI* | *p* |
| (Intercept) | 90.05 | 8.54 – 949.77 | **<0.001** |
| year [2023] | 1.44 | 0.08 – 26.76 | 0.806 |
| chain type [Fast Food & Takeaway] | 0.03 | 0.00 – 0.46 | **0.011** |
| chain type [Pubs, Bars and Inns] | 0.00 | 0.00 – 0.02 | **<0.001** |
| chain type [Restaurants] | 0.01 | 0.00 – 0.08 | **<0.001** |
| continuous [new/removed] | 0.77 | 0.44 – 1.37 | 0.379 |
| year [2023] × chain type [Fast Food & Takeaway] | 0.78 | 0.04 – 14.81 | 0.870 |
| year [2023] × chain type [Pubs, Bars and Inns] | 0.48 | 0.02 – 10.11 | 0.634 |
| year [2023] × chain type [Restaurants] | 0.64 | 0.03 – 12.35 | 0.768 |
| year [2023] × continuous [new/removed] | 0.53 | 0.25 – 1.12 | 0.097 |
| **Random Effects** | | | |
| $\sigma^2$ | 3.29 | | |
| $\tau_{00\ chain:chain\_type}$ | 1.01 | | |
| $\tau_{00\ chain\_type}$ | 0.00 | | |
| $N_{chain}$ | 14 | | |
| $N_{chain\_type}$ | 4 | | |
| Observations | 1037 | | |
| Marginal $R^2$ / Conditional $R^2$ | 0.592 / NA | | |

**Chains perceived as healthy (change in kcal)**

| | kcal | | |
|---|---|---|---|
| *Predictors* | *Incidence Rate Ratios* | *CI* | *p* |
| (Intercept) | 876.85 | 529.12 – 1453.11 | **<0.001** |
| year [2023] | 0.97 | 0.45 – 2.07 | 0.939 |
| chain type [Fast Food & Takeaway] | 0.98 | 0.81 – 1.18 | 0.826 |
| continuous [new/removed] | 1.25 | 1.06 – 1.49 | **0.010** |
| menu section [Children's Menu] | 0.39 | 0.11 – 1.40 | 0.149 |
| menu section [Drinks] | 0.13 | 0.08 – 0.22 | **<0.001** |
| menu section [Mains] | 0.42 | 0.25 – 0.70 | **0.001** |
| menu section [Starters/Sides] | 0.24 | 0.13 – 0.42 | **<0.001** |
| menu section [Sweets/Dessert] | 0.38 | 0.22 – 0.65 | **<0.001** |
| menu section [Toppings/Extras] | 0.14 | 0.06 – 0.32 | **<0.001** |
| year [2023] × chain type [Fast Food & Takeaway] | 1.18 | 0.90 – 1.54 | 0.225 |
| year [2023] × continuous [new/removed] | 0.82 | 0.65 – 1.04 | 0.103 |
| year [2023] × menu section [Children's Menu] | 1.03 | 0.17 – 6.32 | 0.974 |
| year [2023] × menu section [Drinks] | 0.84 | 0.39 – 1.80 | 0.648 |
| year [2023] × menu section [Mains] | 1.09 | 0.51 – 2.34 | 0.824 |
| year [2023] × menu section [Starters/Sides] | 1.11 | 0.48 – 2.55 | 0.811 |
| year [2023] × menu section [Sweets/Dessert] | 1.05 | 0.47 – 2.33 | 0.902 |
| year [2023] × menu section [Toppings/Extras] | 0.36 | 0.10 – 1.33 | 0.124 |
| **Random Effects** | | | |
| $\sigma^2$ | | | |

| | |
|---|---|
| $\tau_{00\ chain:chain\_type}$ | 0.00 |
| $\tau_{00\ chain\_type}$ | 0.00 |
| $N_{chain}$ | 3 |
| $N_{chain\_type}$ | 2 |
| Observations | 904 |

**Chains perceived as healthy (prevalence of mains < 600 kcal)**

| | under600 | | |
|---|---|---|---|
| *Predictors* | *Odds Ratios* | *CI* | *p* |
| (Intercept) | 7.52 | 3.83 – 14.75 | **<0.001** |
| year [2023] | 0.69 | 0.28 – 1.72 | 0.422 |
| chain type [Fast Food & Takeaway] | 1.02 | 0.32 – 3.23 | 0.974 |
| continuous [new/removed] | 0.95 | 0.40 – 2.28 | 0.911 |
| year [2023] × chain type [Fast Food & Takeaway] | 340888403.03 | 0.00 – Inf | 0.998 |
| year [2023] × continuous [new/removed] | 1.51 | 0.43 – 5.30 | 0.524 |
| **Random Effects** | | | |
| $\sigma^2$ | 3.29 | | |
| $\tau_{00\ chain:chain\_type}$ | 0.00 | | |
| $\tau_{00\ chain\_type}$ | 0.00 | | |
| $N_{chain}$ | 3 | | |
| $N_{chain\_type}$ | 2 | | |
| Observations | 377 | | |
| Marginal $R^2$ / Conditional $R^2$ | 0.885 / NA | | |

**Chains presenting themselves as healthy (change in kcal)**

| | kcal | | |
|---|---|---|---|
| *Predictors* | *Incidence Rate Ratios* | *CI* | *p* |
| (Intercept) | 1596.82 | 1299.64 – 1961.96 | **<0.001** |
| year [2023] | 0.92 | 0.78 – 1.09 | 0.347 |
| chain type [Fast Food & Takeaway] | 0.66 | 0.53 – 0.82 | **<0.001** |
| chain type [Pubs, Bars and Inns] | 1.29 | 0.84 – 1.99 | 0.249 |
| chain type [Restaurants] | 0.85 | 0.67 – 1.09 | 0.196 |
| continuous [new/removed] | 1.07 | 1.01 – 1.14 | **0.015** |
| menu section [Children's Menu] | 0.21 | 0.16 – 0.28 | **<0.001** |
| menu section [Drinks] | 0.09 | 0.08 – 0.10 | **<0.001** |
| menu section [Mains] | 0.44 | 0.39 – 0.49 | **<0.001** |
| menu section [Starters/Sides] | 0.23 | 0.20 – 0.26 | **<0.001** |
| menu section [Sweets/Dessert] | 0.29 | 0.25 – 0.33 | **<0.001** |
| menu section [Toppings/Extras] | 0.17 | 0.13 – 0.23 | **<0.001** |
| year [2023] × chain type [Fast Food & Takeaway] | 1.05 | 0.95 – 1.16 | 0.323 |
| year [2023] × chain type [Pubs, Bars and Inns] | 1.12 | 0.89 – 1.40 | 0.327 |
| year [2023] × chain type [Restaurants] | 1.14 | 1.01 – 1.28 | **0.038** |
| year [2023] × continuous [new/removed] | 0.94 | 0.86 – 1.01 | 0.099 |
| year [2023] × menu section [Children's Menu] | 0.97 | 0.68 – 1.38 | 0.853 |
| year [2023] × menu section [Drinks] | 1.01 | 0.85 – 1.20 | 0.887 |
| year [2023] × menu section [Mains] | 1.01 | 0.86 – 1.17 | 0.939 |
| year [2023] × menu | 0.99 | 0.83 – 1.19 | 0.918 |

| | | | |
|---|---|---|---|
| section [Starters/Sides] | | | |
| year [2023] × menu section [Sweets/Dessert] | 1.11 | 0.92 – 1.34 | 0.279 |
| year [2023] × menu section [Toppings/Extras] | 0.98 | 0.69 – 1.38 | 0.896 |
| **Random Effects** | | | |
| $\sigma^2$ | 0.47 | | |
| $\tau_{00\ chain:chain\_type}$ | 0.07 | | |
| $\tau_{00\ chain\_type}$ | 0.00 | | |
| ICC | 0.13 | | |
| $N_{chain}$ | 43 | | |
| $N_{chain\_type}$ | 4 | | |
| Observations | 6575 | | |
| Marginal $R^2$ / Conditional $R^2$ | 0.457 / 0.530 | | |

**Chains presenting themselves as healthy (prevalence of mains < 600 kcal)**

| | under600 | | |
|---|---|---|---|
| *Predictors* | *Odds Ratios* | *CI* | *p* |
| (Intercept) | 4.10 | 1.77 – 9.48 | **0.001** |
| year [2023] | 1.22 | 0.79 – 1.89 | 0.361 |
| chain type [Fast Food & Takeaway] | 0.72 | 0.27 – 1.94 | 0.514 |
| chain type [Pubs, Bars and Inns] | 0.03 | 0.00 – 0.21 | **0.001** |
| chain type [Restaurants] | 0.25 | 0.08 – 0.72 | **0.010** |
| continuous [new/removed] | 0.69 | 0.53 – 0.89 | **0.004** |
| year [2023] × chain type [Fast Food & Takeaway] | 1.04 | 0.65 – 1.66 | 0.879 |
| year [2023] × chain type [Pubs, Bars and Inns] | 0.14 | 0.03 – 0.69 | **0.016** |
| year [2023] × chain type [Restaurants] | 0.62 | 0.37 – 1.05 | 0.074 |
| year [2023] × continuous [new/removed] | 1.31 | 0.92 – 1.86 | 0.131 |
| **Random Effects** | | | |
| $\sigma^2$ | 3.29 | | |
| $\tau_{00\ chain:chain\_type}$ | 1.22 | | |
| $\tau_{00\ chain\_type}$ | 0.00 | | |
| ICC | 0.27 | | |
| $N_{chain}$ | 42 | | |
| $N_{chain\_type}$ | 4 | | |
| Observations | 3251 | | |
| Marginal $R^2$ / Conditional $R^2$ | 0.184 / 0.404 | | |
