## Supplementary Material 3 for "How have restaurant menus changed following England’s calorie labelling regulations and who is likely to benefit? A longitudinal analysis of online menu data"

### Supplementary Material 3: Forest plots of calorie content change estimates

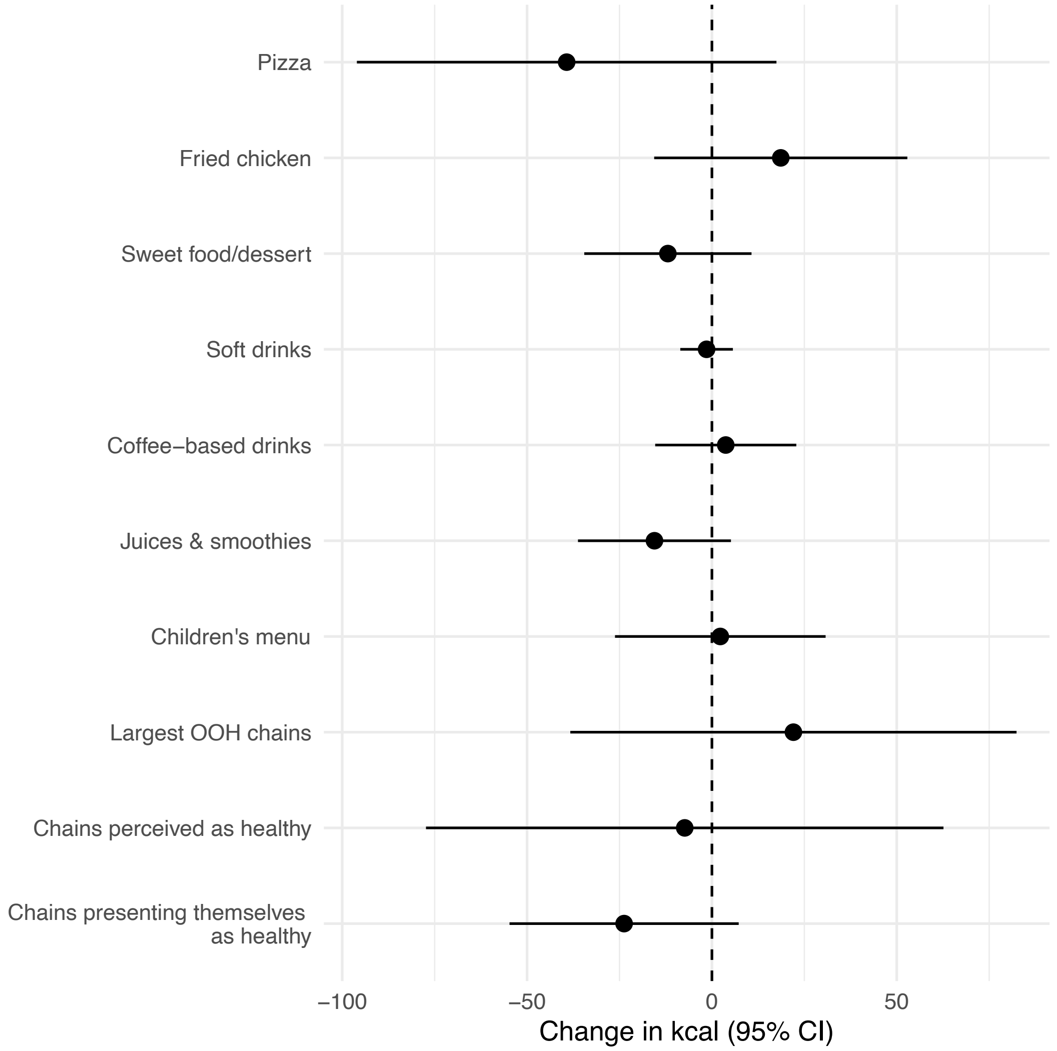


**Figure S2**. Overall estimates of change in calorie (kcal) content between June 2022 and June 2023

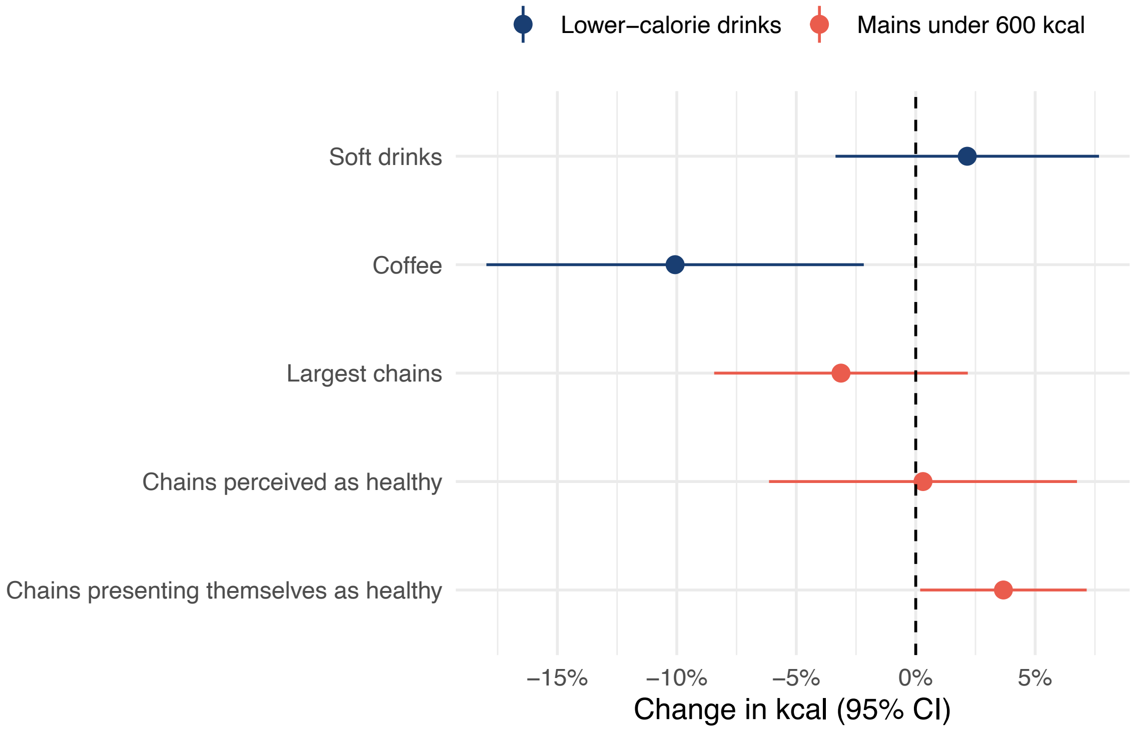


**Figure S3**. Overall estimates of change in % of lower-calorie drinks and mains under 600 kcal between June 2022 and June 202

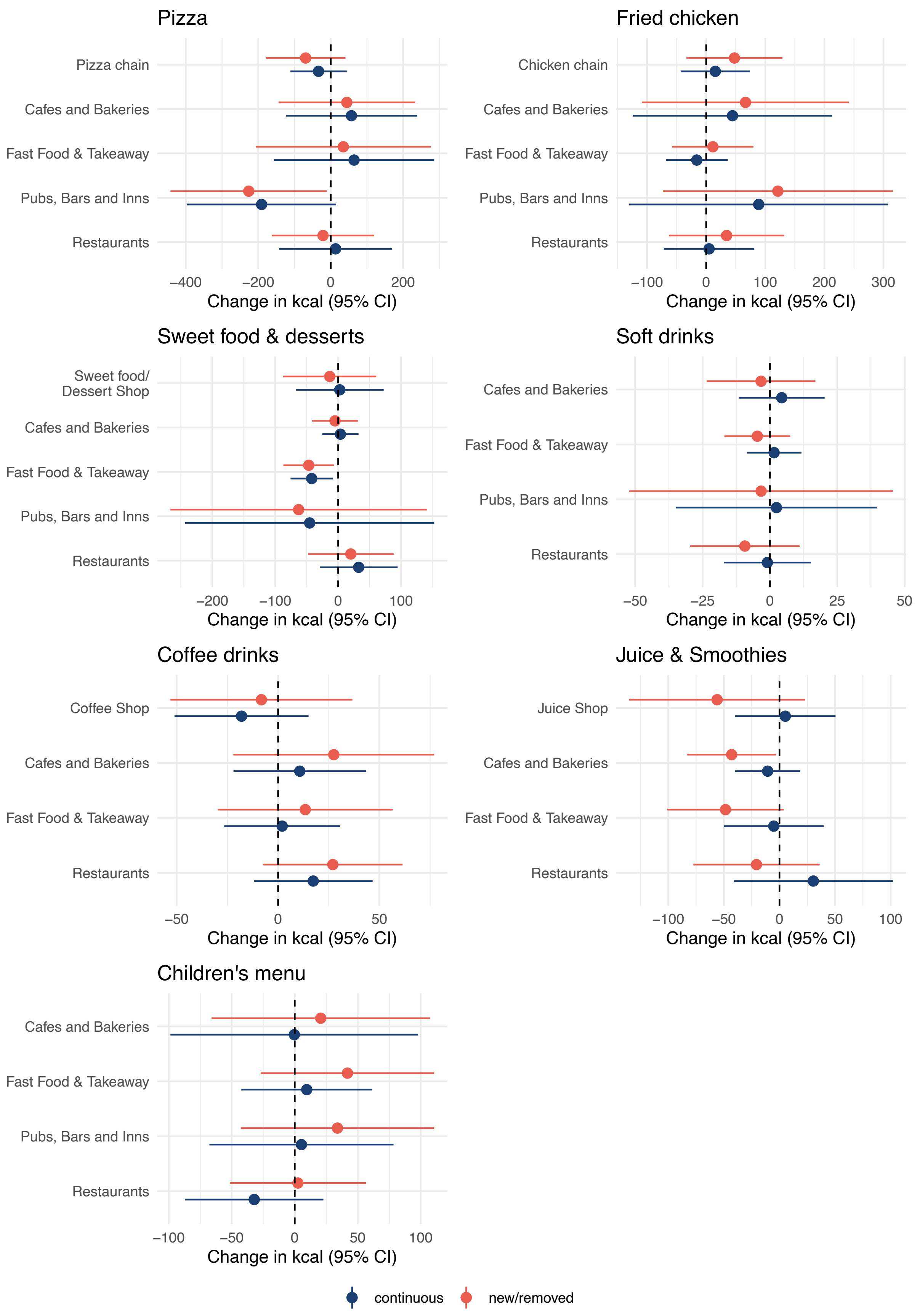


**Figure S4**. Estimates of change in calorie (kcal) content between June 2022 and June 2023 in specific food and drink items by chain type and whether the item was continuously on the menu

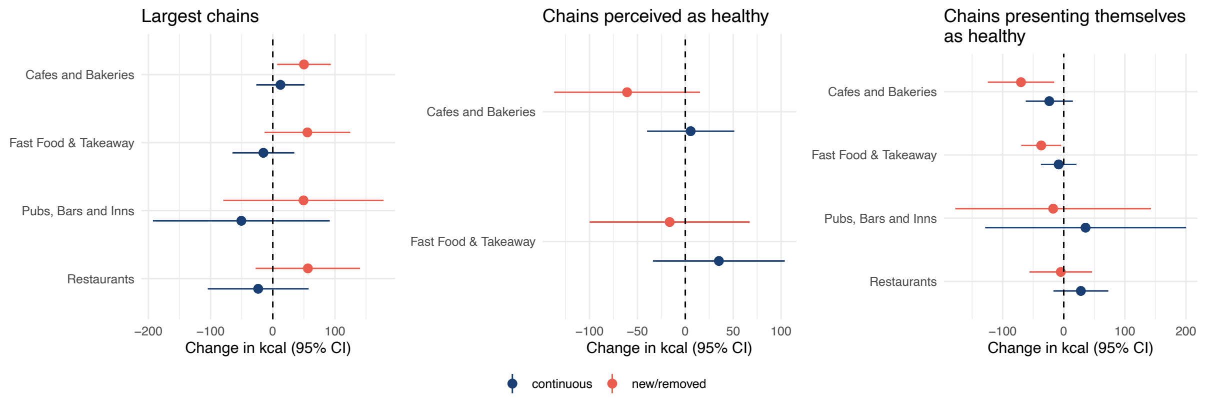


**Figure S5**. Estimates of change in calorie (kcal) content between June 2022 and June 2023 in specific chains by chain type and whether the item was continuously on the menu

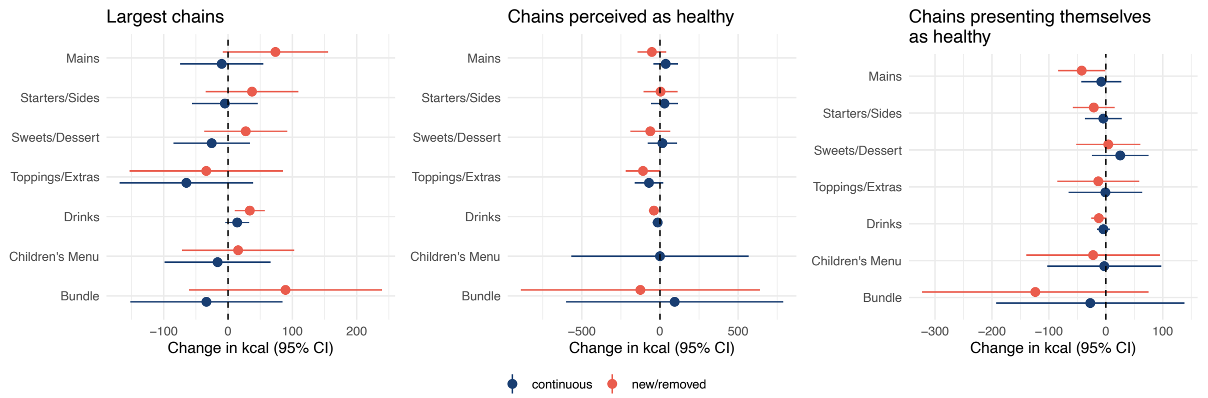


**Figure S6**. Estimates of change in calorie (kcal) content between June 2022 and June 2023 in specific chains by menu section and whether the item was continuously on the menu

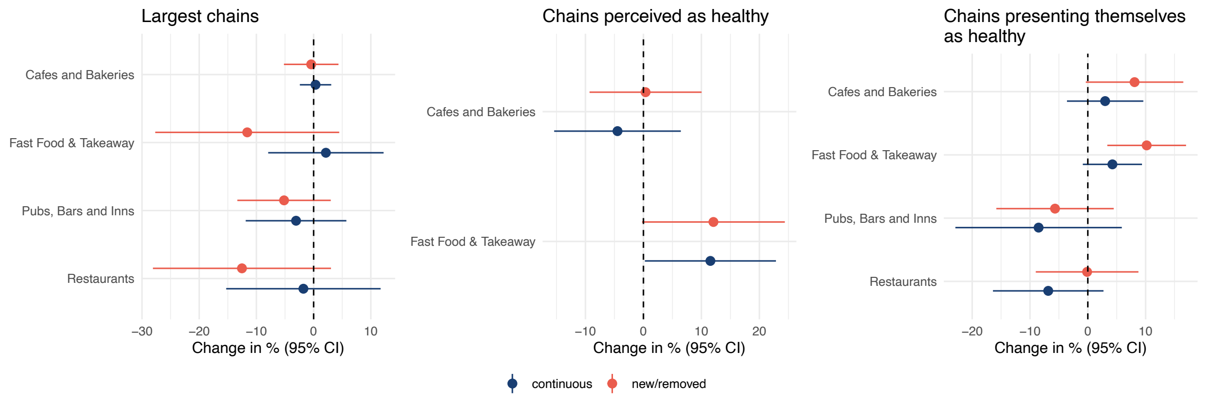


**Figure S7**. Estimates of change in percentage points of main items under 600 kcal between June 2022 and June 2023 in specific chains by chain type and whether the item was continuously on the menu
