## Supplementary Material 4 for "How have restaurant menus changed following England’s calorie labelling regulations and who is likely to benefit? A longitudinal analysis of online menu data"

### Supplementary Material 4: Results from purchasing analysis

*Table S1. Odds ratios (95% CI) of purchasing frequency by sociodemographic characteristics*

| *Case study* | Purchasing frequency | Male vs female | Middle vs high SES | Low vs high SES | Age <35 vs 35–54 years | Age 55+ vs 35–54 years | | | |
|---|---|---|---|---|---|---|---|---|---|
| *Pizza – from pub chains* | Absolute | **1.91 (1.09 to 3.43)** | 1.57 (0.75 to 3.19) | 1.97 (0.82 to 4.83) | 0.74 (0.33 to 1.71) | **2.28 (1.26 to 4.18)** | | | |
| | Relative | 1.32 (0.79 to 2.21) | 1.12 (0.58 to 2.17) | 1.46 (0.66 to 3.26) | 1.11 (0.51 to 2.40) | 1.10 (0.64 to 1.91) | | | |
| | Purchasing frequency | Male vs female | Middle vs high SES | Low vs high SES | Age <25 vs 35–44 years | Age 25–34 vs 35–44 years | Age 45–54 vs 35–44 years | Age 55–64 vs 35–44 years | Age 65+ vs 35–44 years |
| *Pizza – from any large chain* | Absolute | **1.26 (1.07 to 1.49)** | 1.04 (0.84 to 1.27) | **1.33 (1.03 to 1.73)** | 1.24 (0.86 to 1.81) | 1.25 (0.94 to 1.67) | 1.01 (0.80 to 1.28) | 1.05 (0.82 to 1.35) | **0.69 (0.53 to 0.91)** |
| | Relative | 1.16 (0.99 to 1.35) | 1.02 (0.85 to 1.24) | 1.16 (0.92 to 1.47) | 0.99 (0.70 to 1.41) | 1.14 (0.88 to 1.49) | **0.73 (0.59 to 0.91)** | **0.56 (0.45 to 0.71)** | **0.29 (0.23 to 0.38)** |
| *Sweet food/dessert – from Fast-Food/Takeaway* | Absolute | 0.82 (0.62 to 1.09) | 1.23 (0.87 to 1.73) | 0.89 (0.57 to 1.39) | 1.20 (0.67 to 2.22) | 1.39 (0.88 to 2.21) | 0.98 (0.66 to 1.44) | 0.71 (0.46 to 1.08) | **0.62 (0.39 to 0.99)** |
| | Relative | **0.61 (0.46 to 0.81)** | 1.11 (0.80 to 1.55) | 0.83 (0.53 to 1.28) | 1.49 (0.83 to 2.66) | 1.14 (0.73 to 1.77) | 0.79 (0.54 to 1.14) | **0.46 (0.30 to 0.69)** | **0.32 (0.2. to 0.50)** |
| *Sweet food/dessert – from any large chain* | Absolute | 1.07 (0.92 to 1.25) | 1.07 (0.89 to 1.29) | **0.73 (0.57 to 0.92)** | 1.01 (0.72 to 1.44) | **1.51 (1.16 to 1.98)** | 1.12 (0.90 to 1.39) | **1.43 (1.14 to 1.80)** | **2.30 (1.81 to 2.94)** |
| | Relative | **0.82 (0.73 to 0.93)** | 0.95 (0.82 to 1.10) | 0.82 (0.68 to 0.99) | 0.99 (0.73 to 1.33) | 1.01 (0.81 to 1.26) | 0.90 (0.75 to 1.08) | 0.85 (0.71 to 1.02) | 0.86 (0.72 to 1.04) |

| | | | | | | | | | |
|---|---|---|---|---|---|---|---|---|---|
| *Coffee/coffee-based drinks from any chain* | Absolute | **1.30 (1.14 to 1.48)** | 0.98 (0.84 to 1.15) | 1.01 (0.83 to 1.24) | 0.81 (0.61 to 1.10) | **1.29 (1.03 to 1.62)** | **1.33 (1.11 to 1.59)** | **1.69 (1.40 to 2.05)** | **2.13 (1.74 to 2.61)** |
| | Relative | 1.07 (0.97 to 1.17) | 0.96 (0.86 to 1.08) | 0.90 (0.78 to 1.04) | **0.77 (0.62 to 0.97)** | 0.97 (0.82 to 1.15) | 1.03 (0.90 to 1.18) | 1.05 (0.91 to 1.20) | 0.88 (0.76 to 1.02) |
| *Juice/smoothie – from any large chain* | Absolute | 0.89 (0.70 to 1.15) | 1.25 (0.92 to 1.69) | 1.15 (0.78 to 1.69) | 0.81 (0.47 to 1.45) | 0.72 (0.46 to 1.12) | 1.31 (0.92 to 1.85) | 1.00 (0.69 to 1.44) | **1.60 (1.10 to 2.35)** |
| | Relative | **0.78 (0.61 to 0.99)** | 1.19 (0.88 to 1.60) | 1.14 (0.79 to 1.65) | 0.91 (0.51 to 1.63) | 0.84 (0.54 to 1.31) | 0.96 (0.69 to 1.35) | **0.59 (0.41 to 0.84)** | **0.61 (0.42 to 0.87)** |
| *Largest chains* | Absolute | **1.13 (1.04 to 1.23)** | 1.02 (0.92 to 1.14) | 0.91 (0.80 to 1.04) | 1.09 (0.90 to 1.33) | **1.19 (1.03 to 1.39)** | **1.30 (1.15 to 1.47)** | **1.40 (1.23 to 1.59)** | **1.56 (1.37 to 1.79)** |
| | Relative | 0.95 (0.90 to 1.01) | 1.05 (0.98 to 1.13) | 1.03 (0.94 to 1.13) | 1.04 (0.91 to 1.20) | 0.95 (0.86 to 1.05) | **0.88 (0.81 to 0.96)** | **0.70 (0.65 to 0.77)** | **0.53 (0.49 to 0.58)** |
| *Chains presenting themselves as healthy* | Absolute | **1.19 (1.02 to 1.40)** | 0.90 (0.74 to 1.09) | **0.63 (0.49 to 0.81)** | **0.55 (0.38 to 0.79)** | **0.68 (0.52 to 0.90)** | **0.77 (0.62 to 0.96)** | 0.89 (0.70 to 1.13) | **0.67 (0.52 to 0.86)** |
| | Relative | 1.07 (0.93 to 1.23) | 0.91 (0.77 to 1.07) | **0.77 (0.62 to 0.95)** | 0.83 (0.60 to 1.14) | 0.79 (0.62 to 1.00) | **0.74 (0.61 to 0.90)** | **0.54 (0.44 to 0.66)** | **0.42 (0.34 to 0.52)** |

95% CI = 95% confidence interval; SES = occupation-based socio-demographic status

Absolute purchasing frequency defined as total number of purchasing occasions relating to the specific case study reported by the individual between 3rd January and 27th November 2022. Relative purchasing frequency was defined as the share of purchasing relating to the specific case study out of the total number of purchasing occasions reported by the individual during the same period of time. Estimates were retrieved from negative binomial models adjusted for sex, SES (high/middle/low), and age category (<25, 35–44, 45–54, 55–64, 65+ years). Note that for the frequency of pizzas purchased from pub chains, age was included in three categories (<35, 35–54, 55+ years) due to low numbers of observations (n=93/1.5% who reported purchasing pizza from pubs). Authors' analysis of Worldpanel by Numerator's OOH Purchase panel, 47w/e, 27th Nov 2022.
