## Supplementary Material 5 for "How have restaurant menus changed following England’s calorie labelling regulations and who is likely to benefit? A longitudinal analysis of online menu data"

### Supplementary Material 5: Sensitivity analysis

The tables below provide an overview of the menu changes from the sensitivity analysis which excludes items for which multiple calorie values were present in the menu data.

*Table S2. Change in calorie (kcal) content between June 2022 and June 2023 – Sensitivity analysis*

| Category | # items (2023) | # chains (2023) | Unadjusted estimate | | | | Adjusted estimate | | | |
|---|---|---|---|---|---|---|---|---|---|---|
| | | | Mean kcal 2022 | Mean kcal 2023 | Change in kcal (95% CI) | Change in % (95% CI) | Mean kcal 2022 | Mean kcal 2023 | Change in kcal (95% CI) | Change in % (95% CI) |
| Pizza | 317 | 29 | 1,140 | 1,183 | 43.8 (-44.9 to 132.5) | 3.8 (-3.9 to 11.6) | 1,050 | 1,021 | -29.0 (-96.0 to 37.9) | -2.8 (-10.1 to 3.3) |
| Fried chicken | 1,119 | 112 | 825 | 854 | 28.9 (-33.4 to 91.1) | 3.5 (-4.0 to 11.0) | 824 | 837 | 13.4 (-22.7 to 49.5) | 1.6 (-3.0 to 5.5) |
| Desserts | 1,226 | 130 | 638 | 607 | -31.7 (-96.8 to 33.3) | -5.0 (-15.2 to 5.2) | 584 | 568 | -15.2 (-39.1 to 8.7) | -2.6 (-7.5 to 1.3) |
| Soft drinks | 541 | 80 | 89.9 | 78.4 | -11.4 (-22.5 to -0.4) | -12.7 (-25.0 to -0.4) | 80.8 | 78.6 | -2.2 (-9.4 to 5.0) | -2.7 (-13.2 to 5.6) |
| Coffee & coffee-based drinks | 247 | 33 | 152 | 168 | 15.4 (-7.8 to 25.3) | 10.1 (-5.1 to 25.3) | 151 | 152 | 1.1 (-18.0 to 20.3) | 0.7 (-14.1 to 11.6) |
| Juices & smoothies | 193 | 37 | 177 | 168 | -8.6 (-32.1 to 15.0) | -4.8 (-18.2 to 8.5) | 147 | 132 | -14.6 (-35.3 to 6.1) | -9.9 (-40.7 to 2.9) |
| Children's menu | 174 | 39 | 451 | 460 | 8.9 (-43.5 to 61.4) | 2.0 (-9.7 to 13.6) | 404 | 417 | 12.1 (-20.7 to 45.0) | 3.0 (-5.9 to 9.8) |
| Largest chains [a] | 1,183 | 14 | 499 | 493 | -6.2 (-43.3 to 31.0) | -1.2 (-8.7 to 6.2) | 484 | 499 | 15.5 (-20.8 to 51.7) | 3.2 (-4.9 to 9.5) |
| Chains perceived as healthy [b] | 408 | 3 | 304 | 289 | -15.0 (-43.8 to 13.8) | -4.9 (-14.4 to 4.5) | 303 | 294 | -9.2 (-48.4 to 29.9) | -3.0 (-17.5 to 9.1) |
| Chains presenting themselves as healthy [c] | 2,902 | 43 | 489 | 449 | -39.9 (-66.1 to -13.8) | -8.2 (-13.5 to -2.8) | 477 | 460 | -16.9 (-39.9 to 6.0) | -3.6 (-9.2 to 1.2) |

95% CI = 95% confidence interval.

[a] Largest chains determined through most sales recorded in purchasing data per chain type

[b] chains identified in PPI meeting perceived as healthy

[c] chains used tags 'healthy', 'health' or 'healthy options', here referred to as 'presenting themselves as healthy'

Unadjusted change estimates were retrieved using unpaired t tests. Negative binomial regression models included interaction terms between time and chain type as well as between time and whether the item was continuously on the menu, and adjusted for whether the item was on the children's menu, and was intended for sharing, and accounted for items nested in chains which themselves are nested in the type of chain. The model of sweet food/dessert also controlled for whether the item was vegan, while the model of children's menu items also adjusted for whether the item was a meal bundle. Models of specific chain types (largest chains and healthy chains) were also adjusted for menu sections.

*Table S3. Change in prevalence of lower-calorie drinks and mains under 600 kcal between June 2022 and June 2023 – sensitivity analysis*

| Case study | # items (2023) | | Unadjusted estimate | | | Adjusted estimate | | |
|---|---|---|---|---|---|---|---|---|
| | All items | Lower-kcal/under 600 kcal | Mean % 2022 | Mean % 2023 | Change in % (95% CI) | Mean % 2022 | Mean % 2023 | Change in % (95% CI) |
| % lower-calorie drinks | | | | | | | | |
| Soft drinks | 541 | 166 | 28.8 | 30.7 | 1.9 (-3.7 to 7.5) | 27.5 | 30.0 | 2.5 (-3.3 to 8.2) |
| Coffees | 247 | 35 | 25.3 | 14.2 | -10.1 (-18.8 to -3.4) | 25.8 | 14.1 | -11.7 (-20.0 to -3.5) |
| % mains under 600 kcal from | | | | | | | | |
| Largest chains | 325 | 164 | 55.7 | 50.5 | -5.2 (-12.7 to 2.2) | 54.5 | 48.8 | -5.6 (-11.8 to 5.5) |
| Chains perceived as healthy | 159 | 140 | 87.7 | 88.1 | 0.3 (-7.0 to 7.6) | 87.7 | 88.1 | 0.4 (-6.5 to 7.3) |
| Chains presenting themselves as healthy | 1,395 | 970 | 62.3 | 69.5 | 7.2 (3.6 to 10.8) | 63.8 | 67.5 | 3.6 (-0.2 to 7.4) |

95% CI = 95% confidence interval.

[a] Largest chains determined through most sales recorded in purchasing data per chain type

[b] chains identified in PPI meeting perceived as healthy

[c] chains used tags 'healthy', 'health' or 'healthy options', here referred to as 'presenting themselves as healthy'

Unadjusted change estimates were retrieved using z tests of proportions. Logistic regression models included interaction terms between time and chain type as well as time and whether the item was continuously on the menu, and accounted for items nested in chains which themselves are nested in the type of chain.
